# Sex- and Menopause-Specific Hormone-Related ECG Signatures and Incident Cardiovascular Risk in UK Biobank

**DOI:** 10.64898/2026.09.11.26362818

**Authors:** Ángela Hernández, Josseline Madrid, Carlos Sánchez, Alejandro Sanz, William J Young, Patricia B Munroe, Ana Mincholé, Julia Ramírez

**Affiliations:** IIS, Aragon Institute of Engineering Research, University of Zaragoza, Zaragoza, Spain; Centro de Investigación Biomédica en Red, Biomateriales, Bioingeniería y Nanomedicina, Zaragoza, Spain; Endocrinology service, Miguel Servet University Hospital, Zaragoza, Spain; Barts Heart Centre, St Bartholomew’s Hospital, Barts Health NHS Trust, London; William Harvey Research Institute, Queen Mary University of London, London, United Kingdom

**Keywords:** electrocardiography, sex differences, sex hormones, menopause, cardiovascular risk, population health

## Abstract

**Aims:** Sex differences in cardiac electrophysiology and cardiovascular (CV) risk are well established, but the extent to which circulating hormonal markers contribute to electrocardiogram (ECG) morphology remains unclear. We investigated whether hormone-related ECG components are associated with incident CV events and whether these associations differ by sex and menopausal status.

**Methods:** In a prospective cohort (N=8,056 UK Biobank participants; median follow-up: 12.8 years), we first evaluated direct associations between sex hormones and incident CV disease. Based on initial risk signals, two-stage analyses focused on premenopausal females and males. Linear regressions evaluated associations between standardized hormonal markers and residualized lead-I ECG parameters after confounders adjustment. Cox proportional hazards models then tested whether the hormone-predicted ECG components were associated with incident CVD events.

**Results:** In premenopausal females (N=980), higher free androgen index (FAI) was positively associated with QRS amplitude (β=19.17µV, p-value=0.038) and the FAI-predicted QRS amplitude component was associated with incident CVD (Hazard Ratio [HR]=1.41, p-value=0.02). In males (N=3,997), FAI was inversely associated with QTc duration (β=-0.95ms, p-value=0.005), and its predicted component with increased CVD risk (HR=1.32, p-value<0.001). Conversely, higher sex hormone-binding globulin (SHBG) prolonged QTc, with its predicted component also conferring risk (HR=1.15, p-value=0.004). Additionally, FAI directly associated with ST segment deviation (β=19.17µV), whose predicted component was protective (HR=0.76, p-value<0.001).

**Conclusion:** This study shows that SHBG and FAI subtly modulate specific depolarization and repolarization ECG parameters. Relative androgen excess in premenopausal females and lower androgenic status in males appear to be associated with subclinical electrophysiological variations linked to long-term CV risk.

**Graphical Abstract.:** 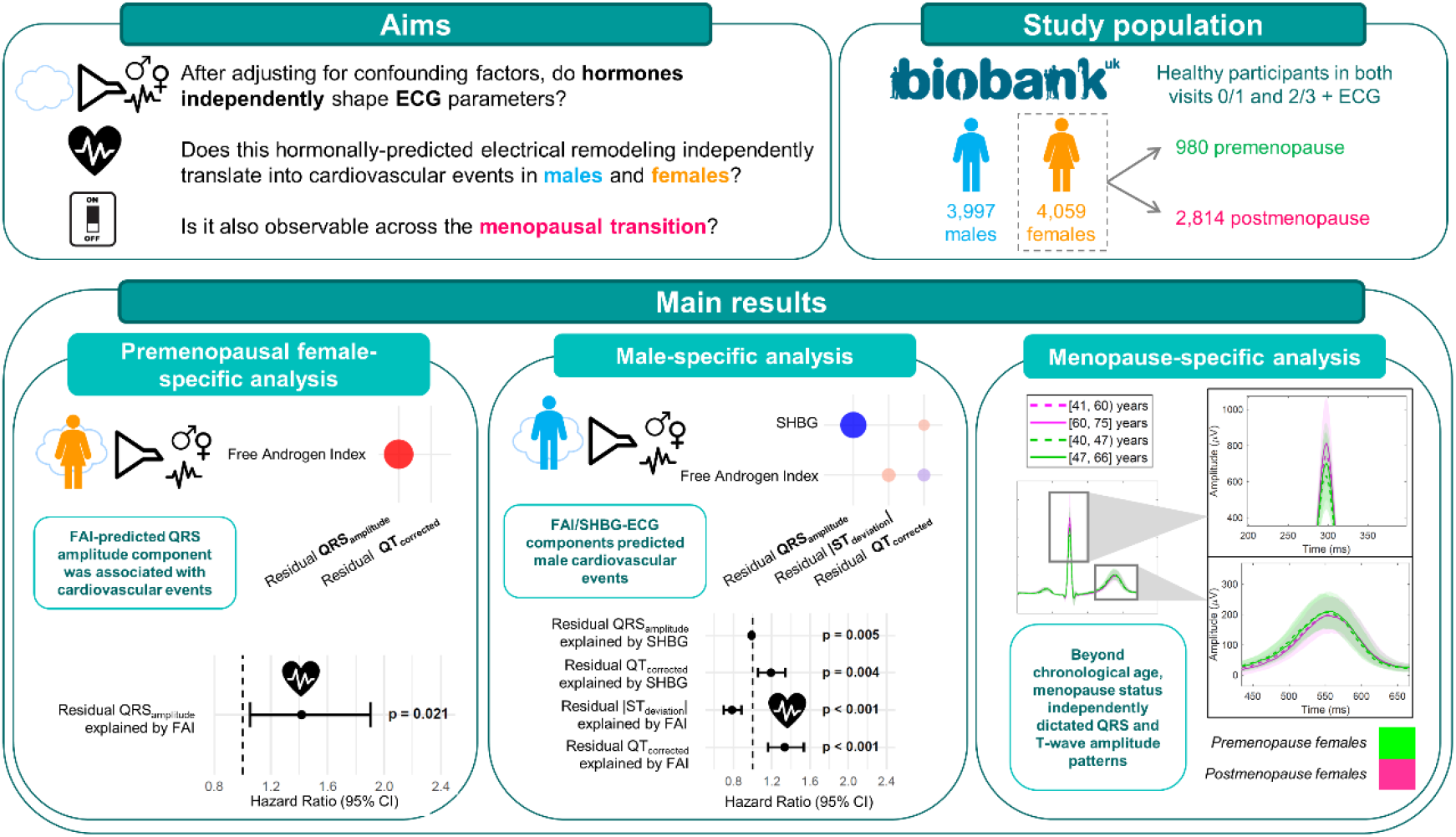

**Lay summary:** This study shows that sex hormone levels are linked to subtle variations in the heart’s electrical activity on an electrocardiogram (ECG) in a middle-aged population without prevalent cardiovascular disease, which may help identify long-term cardiovascular risk in both men and premenopausal women.

- **In premenopausal females:** Higher levels of androgens, specifically those related to free testosterone, were linked to subtle changes in the heart’s electrical signals, specifically higher electrical voltage during heart contraction, and a higher long-term risk of developing cardiovascular disease.
- **In males:** Hormonal balance showed a dual effect; lower active androgen levels were linked to delays in how the heart resets between beats and a higher cardiovascular risk, whereas higher androgen levels appeared protective by supporting a healthier electrical recovery pattern (ST-segment profile) and preventing excessive heart muscle enlargement signals.

## Introduction

Sex differences in cardiac electrophysiological parameters are evident across the surface electrocardiogram (ECG)^1–4^, a widely available non-invasive tool for cardiovascular (CV) phenotyping and risk assessment. Sex-based disparities in CV disease (CVD) prevalence, clinical presentation, and outcomes are also well established^5,6^. However, the relationship between circulating sex hormone concentrations and ECG parameters, and whether hormone-related ECG variation is associated with CV outcomes, remains poorly understood.

Experimental and clinical evidence suggests that sex hormones can modulate ion-channel function and action potential morphology, effects that may be reflected on the surface ECG^1^. For instance, increasing circulating testosterone during male puberty shortens QTc intervals, potentially through enhanced IKs-mediated repolarization and accelerated Ca^2+^ decay, whereas lower androgen levels and oestradiol-mediated inhibition of the rapid delayed rectifier potassium current (IKr) may contribute to the longer QTc intervals observed in females^2^. In addition, ECG morphologies are influenced by structural dimorphism, as females generally exhibit lower left ventricular mass (LVM) and smaller cardiac volumes^7,8^. Recent studies have shown that deep learning models can accurately infer biological sex from the ECG^9^, and discrepancies between ECG-predicted and recorded sex have been linked to hormonal status and CV outcomes^10,11^. However, few large-scale studies have quantified the extent to which circulating hormonal profiles are associated with ECG variation after accounting for cardiac anatomy and clinical factors, or whether these hormone-associated ECG components carry longitudinal clinical relevance.

The menopausal transition marks a major hormonal shift in females, with a marked decline in circulating oestradiol and alterations in the oestrogen-to-androgen balance. This transition is also accompanied by a significantly altered CV physiology^12^ (e.g. dysregulation in glucose homeostasis or arterial hypertension) and increased risk of adverse CVD events^12–14^. Although oestradiol is central to menopausal physiology, androgen-related markers may also influence myocardial electrophysiology^15^ and can be measured at scale in population biobanks^13,14,16^. These include total testosterone, the primary circulating androgen modulating cardiac ion channels^17^; sex hormone-binding globulin (SHBG), the main carrier protein that binds and inactivates free hormones in circulation; and free androgen index (FAI), a more accurate bioavailable testosterone fraction than total testosterone. Nevertheless, whether menopausal endocrine status is reflected in specific ECG phenotypes and whether these phenotypes carry measurable long-term clinical relevance remains unclear.

In this study, we investigated whether hormonal markers are associated with specific ECG parameters beyond anatomical and clinical factors, whether these hormone-associated ECG components differ by sex and menopausal status, and whether these components are associated with incident CV events as a measure of long-term clinical relevance.

## Methods

### Study Population and Exclusion Criteria

The UK Biobank (UKB)^18^ study is a large-scale prospective cohort that contains half a million densely phenotyped individuals from the UK. For the present study, we identified a specific subset of subjects with both ECG recordings from an exercise stress test (EST-UKB; 13-year median follow-up) at visits 0 and 1, and cardiac imaging data from the imaging study (IMG-UKB; 4-year median follow-up) at visits 2 and 3 (Figure 1).

**Figure 1:**
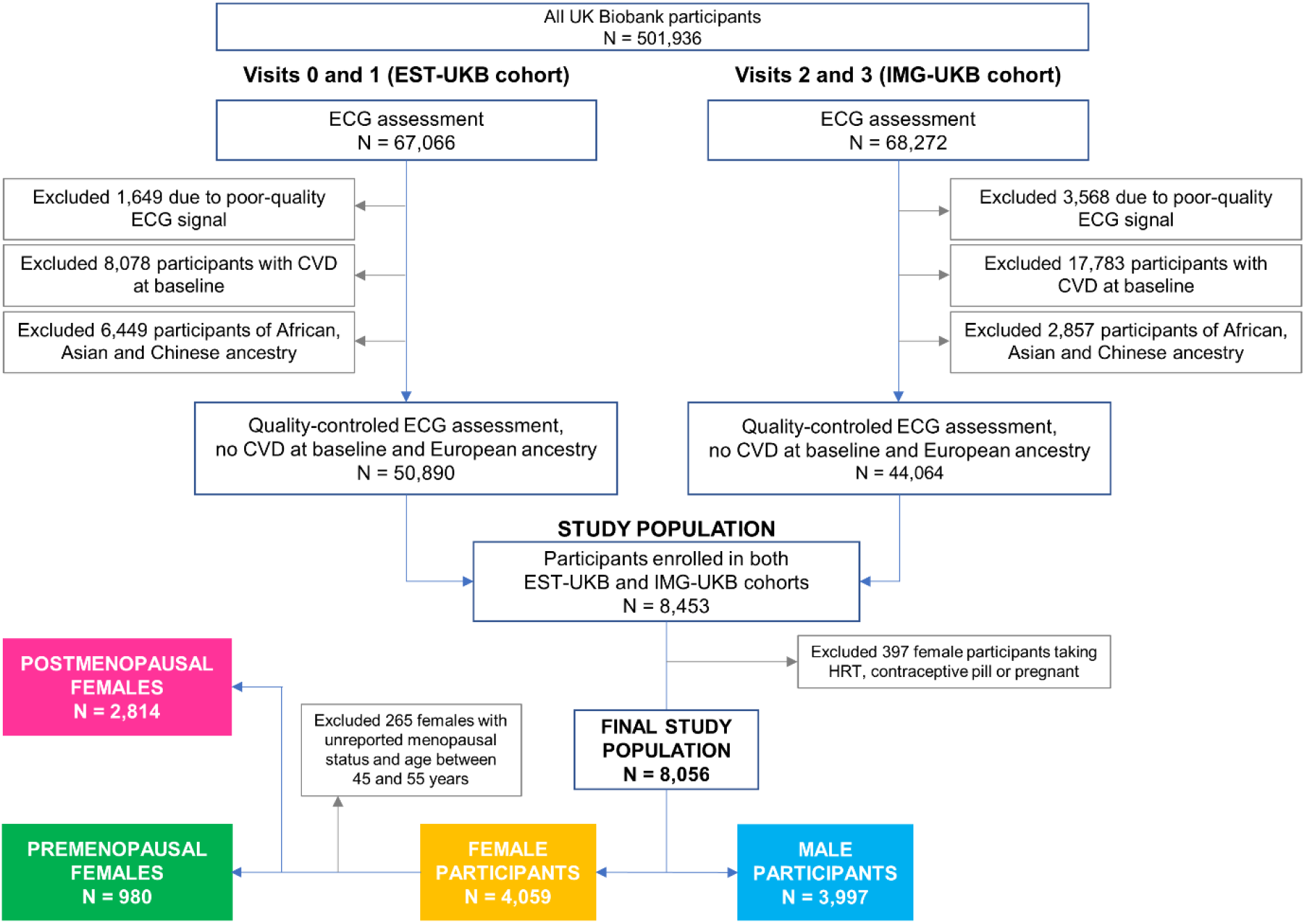
Study population flowchart. Flowchart showing the selection of UK Biobank participants with available lead I resting ECG data from the exercise stress test, hormonal measurements from visits 0/1, and 10-second resting ECG along with cardiac magnetic resonance-derived anatomical variables from visits 2/3. Baseline refers to the date of ECG recording used for the main analyses. CVD: cardiovascular disease, HRT: hormone replacement therapy.

The study population was restricted to individuals of European ancestry to minimize ancestry-related differences^19^. We further excluded participants with prevalent CVD, those receiving beta-blockers or calcium channel blockers (ATC codes C07 and C08), and individuals with poor-quality ECG signals as defined in a previous work^20^. Detailed definitions for CVD cases (classified as prevalent by occurrence before baseline ECG) are provided in the Supplemental Methods.

Participants were stratified by sex, and females were further classified by menopause status, based on self-reported data. A reclassification strategy was applied to females with unreported menopausal status (13.5%): females aged <45 years with measured oestradiol were classified as premenopausal, while those aged >55 years with missing oestradiol were classified as postmenopausal; the remainder were excluded. We also excluded individuals with discordant sex information (discrepancy between self-reported sex and genetic sex). Finally, to ensure a stable hormonal profile, we also excluded female participants who were pregnant or reported using hormone replacement therapy (HRT) or oral contraceptives at the time of assessment. Consequently, the final study population includes 8,056 participants (Figure 1).

General ethical approval was granted for UK Biobank studies by the United Kingdom’s National Health Service Research Ethics Service (11/NW/0382). Participants provided written informed consent for their data to be stored and used for research purposes. This study was conducted under UK Biobank Application Number 8256.

### Selection of Confounding Factors

The following set of confounding factors was considered based on their established influence on cardiac electrophysiology and ECG morphology^21–23^, as well as their documented role in CV risk^24^. Socio-demographic, anthropometric and lifestyle confounders included age, body composition (body mass index (BMI), waist circumference, and fat-free mass), smoking status, alcohol consumption, physical activity levels quantified by the International Physical Activity Questionnaire, and hand grip strength as a proxy for muscular fitness. Selected clinical and metabolic confounders included blood pressure, heart rate, diabetes diagnosis, total cholesterol, and low- and high-density lipoprotein cholesterol (LDL and HDL, respectively).

To account for cardiac anatomical influences on the ECG, we included measures of LVM (indexed to body surface area^7^ using the DuBois formula^25^), wall thickness (LVWT) and ejection fraction (LVEF) as confounders; however, they were exclusively available for the subset of participants who attended the imaging assessment (visit 2 and 3). To account for the time interval between ECG recording visit and cardiac imaging (Δt), an additional interaction term between each anatomical variable and Δt was included in our models as a confounding factor. Detailed descriptions of the UKB field IDs are provided in the Supplemental Methods.

### Selection and Assessment of Hormonal Profiles

Hormonal profiles were derived from measured serum concentrations collected during visits 0 and 1, including total testosterone (nmol/L), oestradiol (pmol/L), and SHBG (nmol/L). From these, the oestradiol/testosterone (O/T) ratio and the FAI (%) were derived. Quality control was performed by applying physiological hard cutoffs to exclude extreme outliers. Specifically, total testosterone measurements exceeding 50 nmol/L in males and 5 nmol/L in females, as well as SHBG measurements exceeding 300 nmol/L in both sexes, were removed. These thresholds were established to mitigate potential assay artifacts, biologically implausible values, or potentially undisclosed exogenous hormone administration (e.g., active testosterone replacement therapy). Detailed descriptions of laboratory assays, UKB field IDs and specific formulas are provided in the Supplemental Methods.

To address missingness, we used Multiple Imputation by Chained Equations (MICE)^26^ with a CART-based approach, excluding variables with more than 20% missing data^27^.

### ECG Pre-processing

After applying participant exclusion criteria, all subsequent analyses were conducted using ECG recordings and confounding factors from visit 0 or 1 (prioritizing the earliest available recording), as serum samples for hormonal profiling were only collected at these visits, enabling the accurate integration of hormonal factors into the models. The ECG signals from visits 0 and 1 were acquired during a bicycle exercise stress test comprising three stages: a resting phase (15 seconds), an exercise phase (6 minutes), and a recovery phase (1 minute)^18^. For this study, we utilized the Lead I recording and exclusively selected the ECG segment from the initial resting stage.

ECG pre-processing involved high-pass filtering at 0.3 Hz, baseline wander correction through cubic splines interpolation, high-frequency noise removal by low-pass filtering at 40 Hz, and exclusion of ectopic beats^28^. A median heartbeat ECG was derived for each participant; from these individual templates, a wavelet-based delineator located ECG wave onset, peaks, and end timings^29^. Characterization of ECG waveforms included standard and morphology-based indices. QRS and T-wave morphologies were quantified using Hermite functions to capture inter-subject variability^20^. In addition, group-level median heartbeats with confidence intervals (CI) were constructed, stratified by sex, age and RR interval. Individual median beats were aligned using cross-correlation, and only those maintaining high morphological consistency (exceeding r = 0.80 correlation coefficient threshold) were included in the final group median calculation. All processing of the ECG signals was performed using custom software in MATLAB R2023a (The MathWorks Inc.)

### Assessment of ECG-derived parameters

Electrophysiological parameters were extracted from the previously generated individual median heartbeats to ensure robust measurements. The PR interval was measured to assess atrioventricular conduction, while the amplitude and duration of the QRS complex were evaluated to characterize the conduction of ventricular electrical activity. Ventricular repolarization was assessed using several measures, including the absolute value of ST segment deviation, the QT interval and T-peak-to-end (Tpe) interval, both corrected using Bazett’s formula (QTc and Tpec, respectively)^30^, as well as T-wave amplitude and T-wave morphological variations quantified using the TMV index^31^, based on deviations from a normal reference.

### Confounders screening and comparative ECG phenotyping by sex and menopausal status

We assessed the normality of continuous confounders using the Shapiro-Wilk test. Multicollinearity among confounders was evaluated via Spearman (continuous) and Phi (categorical) correlation analyses. For pairs with a correlation coefficient greater than 0.8, one variable was excluded and the most physiologically relevant variable was retained. Following this screening, continuous variables were summarized as median and interquartile range (IQR), and compared across groups (sex and menopausal status) using the Mann-Whitney U test, while categorical variables were reported as counts and percentage, and compared using the Chi-squared test.

Furthermore, the comparative analysis of ECG followed a two-step approach. First, the distributions were visualized using sex- and menopause-stratified boxplots, retaining only those metrics with statistically significant differences between groups for downstream analyses. Second, we constructed median heartbeats with 25^th^-75^th^ interquartile envelopes to highlight sex- and menopause-specific morphological variance in lead I before multivariate adjustment. These heartbeats were further stratified to visualise the influence of aging (minimum age to 55 years and 55 to maximum age). These specific strata were selected based on the data distribution to ensure a balanced and statistically robust representation of data across all subgroups. All statistical tests were two-sided, and statistical significance was defined as p-value≤0.05.

### Association of hormonal markers with incident CVD risk

Univariable Cox proportional hazards models were first performed to assess the association between hormonal markers and incident CVD risk within sex- and menopause-stratified groups. All hormonal measurements were standardized using intra-subgroup z-scores to allow for uniform risk estimates per 1-standard deviation (SD) increment within each respective stratum. This initial phase served as a strict eligibility criteria: only the specific hormone-stratum combinations that demonstrated a statistically significant (p-value≤0.05) Hazard Ratio (HR) for incident CVD were carried forward into downstream electrophysiological mediation analyses.

### Adjustment for measured confounders in ECG parameters and association with CV events

To assess whether the significant hormone markers associate with ECG parameters, a two-step regression framework was applied. First, to estimate ECG variation not attributable to the set of confounders, linear regression models were fitted for each ECG parameter using these confouners as predictors. Continuous predictors were standardised using z-scores to allow for comparison and collinearity was assessed before model fitting. Raw unstandardized residuals, expressed in milliseconds for duration metrics and microvolts for amplitude metrics, were then extracted from these models as covariate-adjusted ECG residuals independent of confounders. Statistical significance was defined as p-value≤0.05.

Secondly, these covariate-adjusted ECG residuals were individually regressed against the corresponding z-score standardized hormonal exposures that exhibited significant association signals in the primary CV risk screening. Linear regression coefficients (β) reflect the net unit change in covariate-adjusted ECG residuals preserving their original units (milliseconds or microvolts) per 1-SD increment in intra-subgroup hormone levels. Then, individual fitted values extracted from these models were obtained as hormone-predicted ECG components, representing the specific fraction of residual ECG variation linearly associated with each hormonal marker under the specified model.

Finally, univariable Cox proportional hazards models were used to evaluate whether these hormone-predicted ECG components were independently associated with incident CV events over a median follow-up of 12.8 years. Participants experiencing incident CVD events within 6 months following baseline ECG were excluded to account for potential undiagnosed cases. HRs and their 95% CIs were calculated per 1-SD increase in each hormone-predicted ECG component. Each fitted hormone-predicted ECG component was entered standardized as the primary predictor to estimate future CV risk, defined as a composite endpoint including major adverse CV events (i.e. heart failure, ischemic heart disease, myocardial infarction and cardiac device implantation), stroke, atrial fibrillation, cardiac disease or self-reported diagnosis. Proportional hazards assumptions were assessed using Schoenfeld residuals. Detailed definitions for CVD cases (classified as incident by occurrence after baseline ECG) are provided in the Supplemental Methods.

## Results

### Study population and variables selection

The final study population included 4,059 (50.4%) females and 3,997 (49.6%) males (Figure 1). Females and males had similar age distributions but differed markedly in anthropometry, cardiac structure, blood pressure and hormonal markers, consistent with sex-specific profiles (Table 1). Among females, 980 (24.15%) were premenopausal and 2,814 (69.33%) postmenopausal (265 (6.5%) excluded). Postmenopausal females were older and had significantly higher blood pressure, lower fat-free mass and lower concentrations of testosterone, oestradiol, SHBG and FAI (Table 1).

**Table 1:**
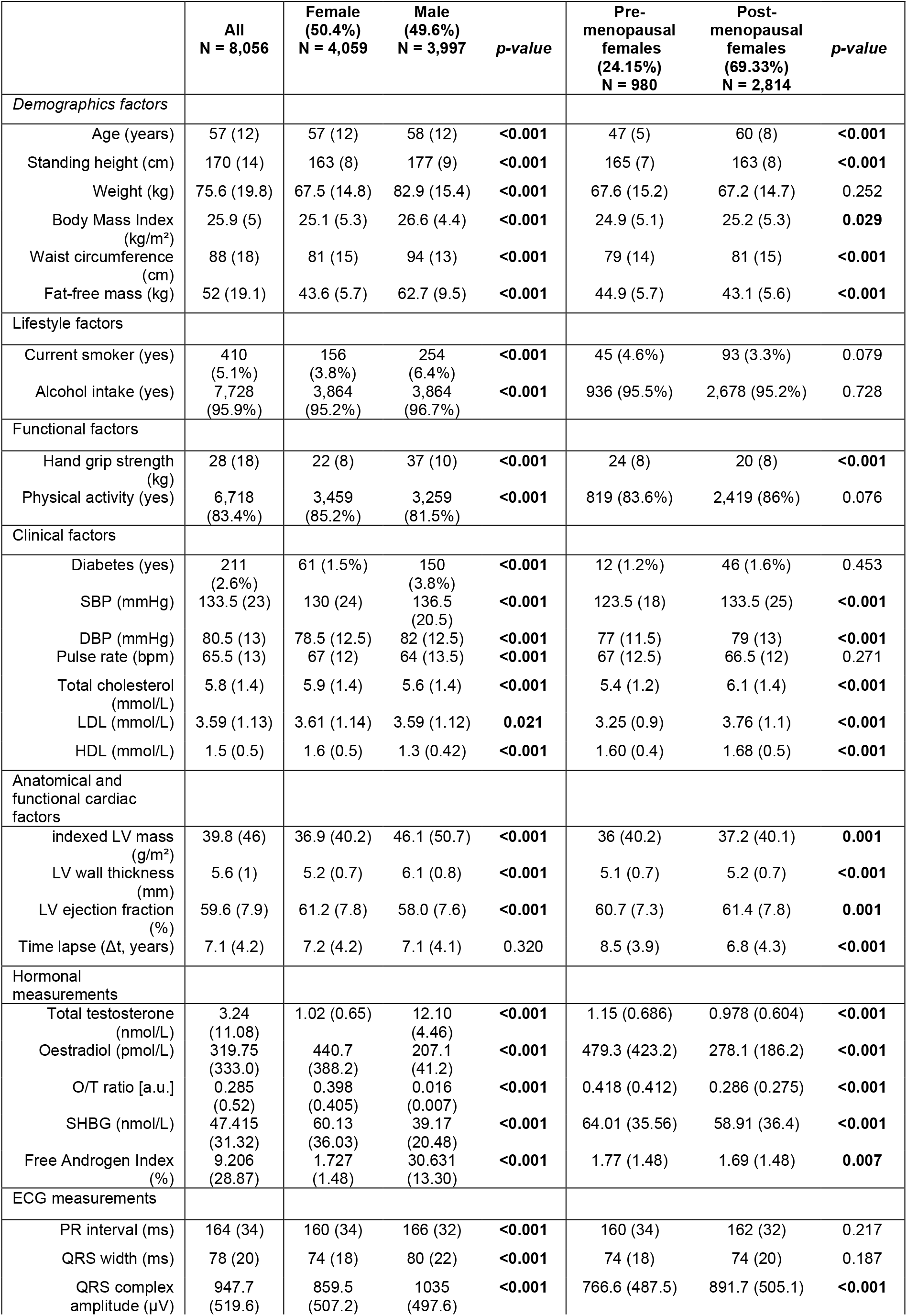
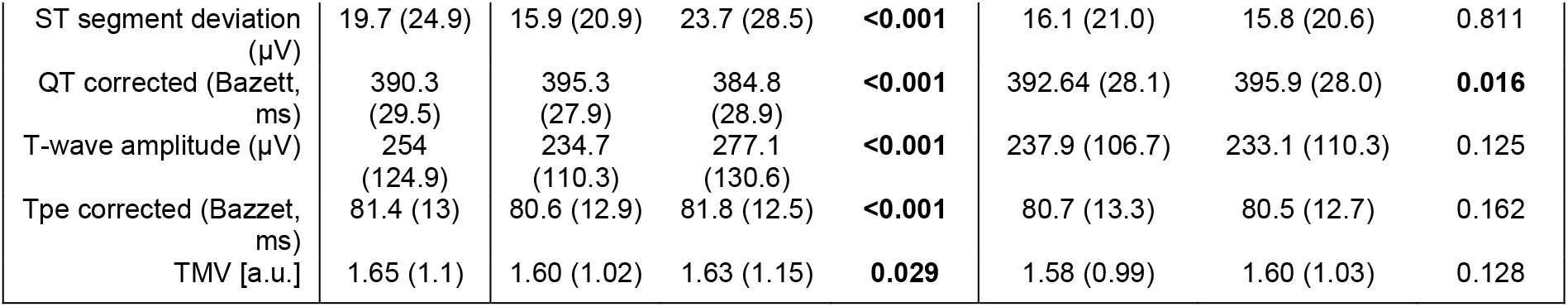
Baseline characteristics of participants in the study, stratified by sex and by menopausal status. Data are presented as median and interquartile range for continuous variables, and as absolute number and percentages for dichotomized variables. All continuous variables were compared using the Mann–Whitney U-test and categorical variables using the Chi-square test. For sex-specific columns, percentage for each count is reported as the percent of that sex subgroup and, for menopause-specific columns, as the percent of that female subgroup. SBP: systolic blood pressure; DBP: diastolic blood pressure; LDL: low-density lipoprotein; HDL: high-density lipoprotein; O/T: Oestradiol/Testosterone; SHBG: sex hormone binding globulin; LV: left ventricle; ECG: electrocardiogram; TMV: T-wave Morphology Variability index. Bold p-values indicate statistical significance at the p-value≤0.05.

All continuous confounders showed a non-normal distribution. Weight, standing height, total cholesterol and LV wall thickness were excluded from subsequent analyses due to high redundancy with BMI, waist circumference and fat-free mass, fat-free mass, LDL and LVEF, respectively (r > 0.8, Supplemental Figure 1), and no high correlation was found between categorical confounders (Supplemental Figure 2). Additionally, oestradiol and, consequently, the O/T ratio, were also excluded due to an excessive number of missing values^32^; specifically, oestradiol data were unavailable for 3,238 women (79.8%) and 3,678 men (92%).

### Sex-specific distribution and morphological characterization of ECG signals

The comparative analysis confirmed previously reported sex differences in electrophysiological parameters^1–4^. As shown in Table 1 and Figure 2 (A.1), females showed longer QTc intervals (395.3 vs. 384.8 ms), whereas males exhibited higher Tpec values (81.8 vs. 80.6 ms), longer PR intervals (166 vs. 160 ms), wider QRS complexes (80 vs. 74 ms), and larger QRS, ST, and T-wave amplitudes (1035 vs. 859.5 µV; 23.7 vs. 15.9 µV; and 277.1 vs. 234.7 µV, respectively) (all p-value<0.001).

**Figure 2:**
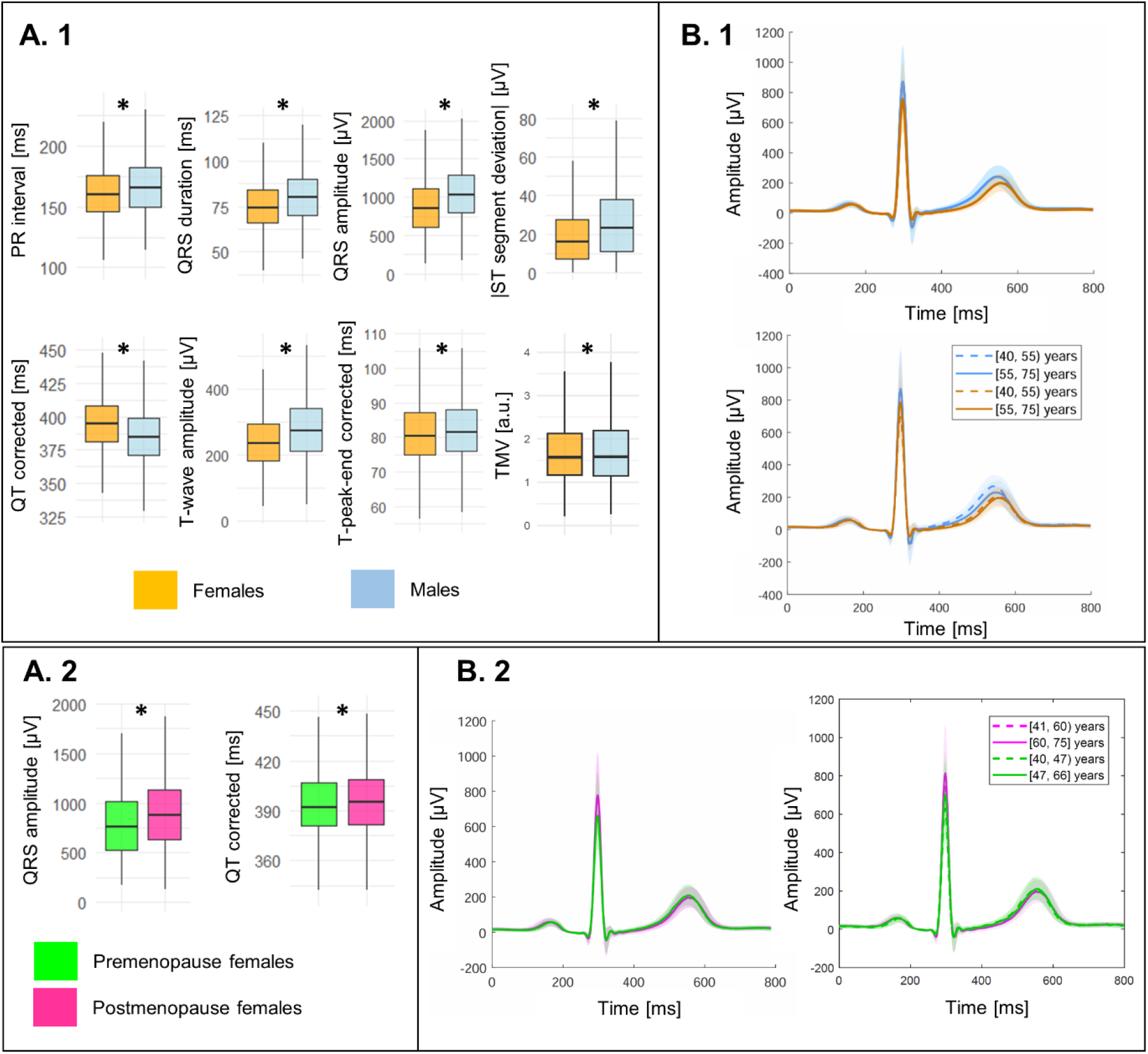
Sex- and menopause-specific distribution and morphology of lead I ECG parameters. (A) Boxplots showing the distribution of ECG intervals, amplitudes, and morphology-derived indices stratified by sex (A.1) and menopausal status (A.2). Boxes show median and interquartile range. Between-group comparisons were performed using the Mann-Whitney U test. (B) Median lead I heartbeat morphology stratified by sex (B.1) and menopause status (B.2). Shaded bands represent the 25^th^-75^th^ percentile range of aligned individual median beats. Morphologies are shown for the total cohort and age strata. High-magnification views of QRS and T-wave morphology are provided in Supplemental Figures 1 and 4. ECG, electrocardiogram; RR, RR interval. *p-value≤0.05.

These differences were also reflected morphologically as shown in Figure 2 (B.1), with females displaying lower amplitudes and a longer QT intervals, whereas males showed higher R-wave peaks and more prominent T-waves (Figure 2 (B.1, above)). Age-stratified analysis (Figure 2 (B.1, below)) demonstrated that the older cohort (55–75 years) exhibited a slight reduction in T-wave amplitude and a broader QTc duration compared to the younger group (40–55 years), while preserving a distinct separation between sexes across age strata. High-magnification details are provided in Supplemental Figure 3.

Table 1 also outlines the baseline characteristics, demonstrating extensive and several statistically significant differences between pre- and postmenopausal cohorts. Regarding electrocardiographic parameters, postmenopausal females were uniquely characterized by significantly higher QRS amplitude (891.6 vs. 764.2 µV; p < 0.001), and slightly longer QTc (396.2 vs. 393 ms, p = 0.013), whereas all other ECG parameters showed no significant differences and were therefore not included in subsequent analyses.

These findings were further corroborated by the boxplots and median ECG morphologies in Figure 2 (A.2 and B.2), respectively. Menopause-stratified visualization (Figure (B.2, left)) showed that postmenopausal females (magenta) were characterized by higher QRS amplitudes and lower, delayed T-wave peaks, compared to premenopausal females (green). Crucially, age-stratified visualization (Figure 2 (B.2, right)) showed that menopause-related morphological differences remained apparent within age strata. Although QRS amplitude increased with age in females, younger postmenopausal females (41–60 years) displayed higher QRS amplitudes than older premenopausal females (47–66 years), supporting a menopause-related ECG signature beyond chronological age. T-wave morphology showed smaller accompanying differences. High-magnification details are provided in Supplemental Figure 4.

### Screening of significant hormonal CVD risk signals across sex and menopause strata

In the initial screening phase, we found that FAI in premenopausal females and both SHBG and FAI in males, were shortlisted to investigate whether their specific CV risk signals were captured by hormone-driven ECG variations (Figure 3). This screening analysis was performed to establish which hormonal markers were associated with incident CVD before evaluating whether these risk signals were reflected in hormone-related ECG components.

**Figure 3.**
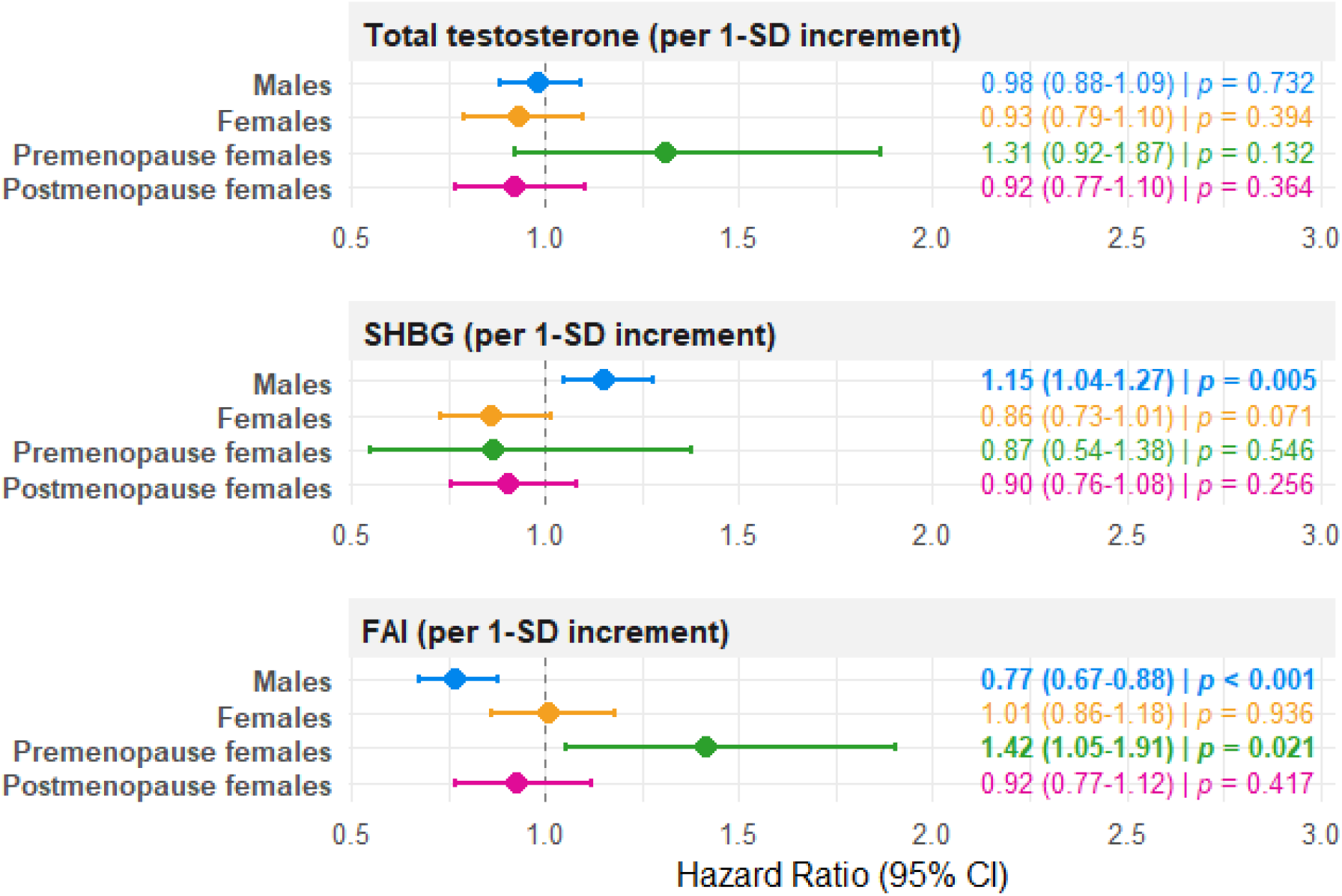
Association between hormone levels and incident cardiovascular disease risk across sex- and menopause-stratified subgroups. Univariable Cox proportional hazards models were evaluated separately in males (blue), overall females (orange), premenopausal females (green), and postmenopausal females (magenta). All hormonal exposures were z-score standardized within each stratum. Point estimates represent the HR (per 1-SD increment) and horizontal error bars correspond to the 95% CIs. Bold p-values indicate statistical significance at the p-value≤0.05 and served as the eligibility criterion for downstream electrophysiological mediation analyses. HR, hazard ratio; CI, confidence interval; SD, standard deviation; SHBG, sex hormone-binding globulin, FAI, free androgen index; p, p-value.

Specifically, among premenopausal females each 1-SD increase in FAI was significantly associated with an elevated risk of incident CVD (HR=1.42 [95% CI: 1.05– 1.91]; p-value=0.021). Conversely, among males each 1-SD increase in SHBG levels was significantly associated with an increased risk (HR=1.15 [95% CI: 1.04–1.27]; p-value=0.005), whereas each 1-SD increase in FAI demonstrated a marked protective effect (HR=0.77 [95% CI: 0.67–0.88], p-value<0.001). In contrast, total testosterone showed no statistically significant associations across any analyzed subgroup, nor did any hormonal marker in postmenopausal females.

### Confounders Adjustment and Determinants of ECG parameters

Focusing on the most relevant findings, confounding analyses revealed distinct electrophysiological modulations between premenopausal females and males (Figure 4, A.1 and A.2, respectively). Across both groups, higher BMI and waist circumference and lower fat-free mass were associated with increased QRS amplitude, while higher fat-free mass and DBP were associated with longer QTc duration. Specifically, in premenopausal females higher BMI was associated with shorter QTc duration, whereas the opposite association was shown in males. Among males, demographic and lifestyle factors exerted a generalized influence across almost all ECG parameters. In contrast, cardiac structural and anatomical metrics concentrated their primary effects on PR interval, QRS amplitude and the absolute value of ST segment deviation. Meanwhile, clinical and metabolic conditions (such as diabetes, pulse rate, and blood pressure) predominantly governed ventricular repolarization dynamics like QTc, T-wave amplitude and Tpec. Individual effect directions, normalized β-coefficients and p-values are provided in Supplemental Table 1 and 2 for premenopausal female and male participants, respectively.

**Figure 4.**
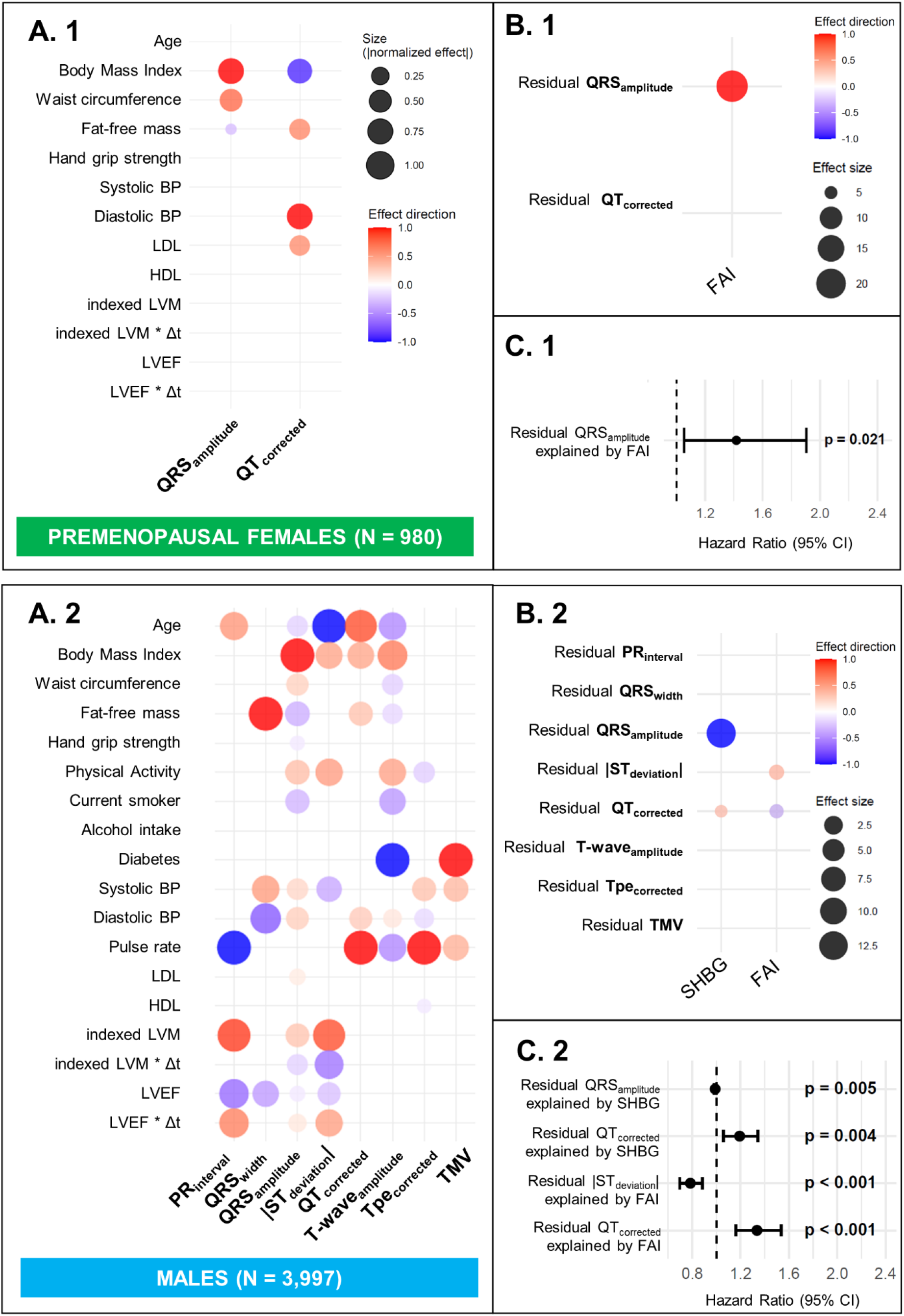
Confounders-adjusted ECG parameters, hormone-related ECG components, and incident cardiovascular events. (A) Standardised associations between measured demographic, functional, lifestyle, clinical, and CMR-derived anatomical confounders and lead I-derived ECG parameters in premenopausal females (A.1) and males (A.2). (B) Associations between hormonal markers and confounders-adjusted ECG residuals in premenopausal females (B.1) and males (B.2). Dot size represents the absolute standardised regression coefficient, and color indicates direction of association. Only significant associations (p-value≤0.05) are displayed in panels A and B. (C) Cox proportional hazards models evaluating the association between hormone-related ECG components and incident cardiovascular events in premenopausal females (C.1) and males (C.2). Hazard ratios are reported per 1-SD increment. BP, blood pressure; CMR, cardiovascular magnetic resonance; FAI, free androgen index; LVM, left ventricular mass; LVWT, left ventricular wall thickness; SHBG, sex hormone-binding globulin; T, testosterone; Δt, time elapsed between hormonal and CMR assessment.

### Association of hormonal factors with ECG Residuals

After adjustment for clinical, anthropometric, lifestyle, and anatomical confounders, ECG residuals were tested for association with hormonal markers. In premenopausal females (Figure 4, B.1), FAI demonstrated a significant positive association with QRS amplitude residuals (β = 19.17 µV per 1-SD increment [95% CI: 1.01 – 37.33]; p-value=0.038). Among males (Figure 4, B.2), distinct electrophysiological profiles emerged for SHBG and FAI: SHBG exhibited an inverse association with QRS amplitude residuals (β = –13.19 µV [95% CI: -23.29 – -3.08]; p-value=0.010), while demonstrated an opposite pattern with a significant direct relationship with QTc residuals (β = 0.83 ms [95% CI: 0.16 – 1.49], p-value=0.014). Meanwhile, FAI was positively associated with ST segment deviation residuals (β = 1.12 µV [95% CI: 0.47 – 1.77]; p-value<0.001) and inversely with QTc residuals (β = –0.95 ms [95% CI: -1.61 – -0.28], p-value=0.005). These results indicate sex- and menopause-specific associations between available hormonal markers and covariate-adjusted ECG components, particularly involving QRS amplitude, ST segment deviation and QTc. Detailed regression estimates, including β-coefficients and p-values, are also provided in Supplemental Table 1 and 2.

### Association Between Hormone-predicted ECG components and CV risk

During a median follow-up of 12.8 years, 20 CV events (2.04%) were reported in the premenopausal female cohort and 335 CV events (8.4%) in the male cohort. No incident CVD events were recorded within 6 months post-ECG baseline. In the premenopausal female cohort (Figure 4, C.1), the FAI-predicted QRS amplitude component was associated with CV risk (HR = 1.417 per 1-SD increment [95% CI: 1.055 – 1.905]; p-value=0.021). In the male cohort (Figure 4, C.2), several hormone-predicted ECG components were significantly associated with incident CVD. The SHBG-predicted QRS amplitude component was associated with a protective effect for incident CV risk (HR = 0.866 per 1-SD increment [95% CI: 0.784 – 0.956]; p-value=0.004), whereas SHBG-predicted QTc component was associated with higher risk (HR = 1.158 per 1-SD increment [95% CI: 1.048 – 1.279]; p-value=0.004). In contrast, the FAI-predicted ST segment deviation component was significantly associated with lower CV risk (HR = 0.760 per 1-SD increment [95% CI: 0.665 – 0.868]; p-value<0.001), while FAI-predicted QTc component was associated with higher risk (HR = 1.317 per 1-SD increment [95% CI: 1.152 – 1.506]; p-value<0.001). Detailed Cox analysis estimates are provided in Supplemental Table 3.

## Discussion

Our findings identify sex- and menopause-dependent associations between hormonal markers and specific lead I-derived ECG components, independently of demographic, anthropometric, and anatomical factors. Specifically, SHBG, and FAI were associated with variations in QRS amplitude, ST segment deviations and the QTc interval, supporting an endocrine contribution to cardiac electrophysiology. Menopause was associated with a distinct ECG remodelling pattern in females, mainly reflected in QRS amplitude beyond chronological age. Importantly, hormone-related ECG components were significantly associated with incident CVD risk in premenopausal females and males, highlighting the potential utility of endocrine-aware ECG phenotyping for CV risk stratification.

A main finding of our study is the potential sex- and menopause-specific endocrine modulation of ECG markers after adjustment for a set of traditional confounders (Figure 4, Panels B1 and B2). In previous literature, experimental animal models^33,34^ and human clinical studies^35,36^ with hormonal interventions have shown that oestradiol and testosterone influence the QTc intervaI^1,17,37^. However, evidence has largely centred on QTc, while relationships between hormonal markers and broader ECG metrics, including depolarization and amplitude-based markers, remain largely unexplored. Here, we extend this focus beyond QTc by assessing hormone-related variation across depolarization, repolarization, and amplitude-based ECG markers.

Another key finding was that the prognostic relevance of hormone-predicted ECG components appeared to be sex- and menopause-specific (Figure 4, Panels C1 and C2). In previous literature, circulating sex hormones and SHBG levels have been established as predictors of clinical outcomes, such as myocardial infarction in both sexes^16^. However, whether hormone-associated ECG components are linked to long-term risk has remained largely unexplored. Here, we show that hormone-predicted ECG components are associated with incident CVD risk in premenopausal females and males, supporting endocrine-aware ECG phenotyping as a promising approach for CV risk stratification.

Among premenopausal females, our findings reveal a plausible relationship linking relative androgen excess to subclinical electrophysiological changes and heightened CV risk. Higher baseline FAI was directly associated with an elevated risk of incident CVD (Figure 3, HR = 1.42, p-value=0.021) and was the sole hormonal marker displaying significant electrophysiological associations in this group, being associated with an increase in QRS amplitude (β= 19.17 µV per 1-SD increment, p-value=0.038) after confounding factors adjustment. Crucially, the FAI-predicted QRS amplitude component was associated with future CV events (HR = 1.417 per 1-SD increment, p-value=0.021). In young and middle-aged females, conditions characterized by androgen excess, such as polycystic ovary syndrome, are known to promote adverse cardiac remodelling^38^. Consistent with this literature, our findings suggest that higher relative androgenicity may also be reflected in subtle ECG-derived depolarization features, potentially reflecting early subclinical ventricular alterations associated with long-term CV risk, even in a lower-risk premenopausal cohort. However, these findings should be interpreted within the context of the low event rate observed in this cohort (20 incident events among 980 premenopausal females; 2.04%), which may have limited statistical power. Furthermore, data on the primary female sex hormones were either insufficient (i.e., oestradiol) or unavailable (i.e., progesterone) for this study, meaning their potential prognostic role remains unknown^32^.

Among males, higher FAI levels were inversely associated with QTc duration (Figure 4 [B.2], β = –0.951 ms), reflecting the well-established physiological role of androgens in shortening ventricular repolarization. Consequently, higher values of the FAI-predicted QTc component capture a relative QTc prolongation associated with a reduced androgenic activity (low FAI). As higher FAI conferred a marked direct protective effect against incident CV events (Figure 3, HR = 0.77 per 1-SD in FAI increment), the positive association between the FAI-predicted QTc component and CV risk (Figure 4 [C.2], HR = 1.317 per 1-SD increment) aligns with previous observation^3^: it shows that subtle QTc prolongation secondary to functional androgen deficiency serves as a sensitive marker of CV risk, potentially reflecting either intrinsic repolarization changes or underlying subclinical ischemic burden.

Similarly, higher SHBG levels, which are consistent with lower free testosterone and reduced androgenic activity, were associated with longer QTc interval (Figure 4 [B.2], β = 0.833 ms). Appropriately, the SHBG-predicted QTc component was also associated with increased CV risk (Figure 4 [C.2], HR = 1.158 per 1-SD increment). Together, the QTc findings from FAI and SHBG support a coherent pattern aligned with existing literature, in which lower androgenic status is linked to prolonged ventricular repolarization and higher CV risk in males^33,35,39,40^. Mechanistically, testosterone enhances repolarizing potassium currents (IKr and IKs) and inhibit L-type calcium currents (ICa,L), thereby shortening the action potential duration.

In addition to ventricular repolarization, androgenic status was associated with the QRS complex and ST-segment deviation in males. FAI was directly associated with ST-segment deviation residuals (β = 1.12 µV per 1-SD increment, p-value<0.001) after confounding factors adjustment, and the resulting FAI-predicted ST segment deviation component showed an inverse association with CV risk (HR = 0.760 per 1-SD increment, p-value<0.001). From a mechanistic perspective, higher androgen status has been associated with the physiological ST-segment elevation typically observed in healthy adult males^41^. Clinical observations in individuals undergoing androgen deprivation therapy demonstrate a significant attenuation of this baseline ST-segment deviation^41^, suggesting that active androgen exposure is required to maintain this male-pattern electrophysiological profile. It is plausible that such androgen-driven baseline elevation provides a physiological buffer that protects against subclinical ST-segment depression, a well-established marker of adverse CV events^16^, although further studies are needed to establish the exact sequence of these associations.

Conversely, SHBG exhibited an inverse association with QRS amplitude residuals (β = –13.19 µV per 1-SD increment, p-value=0.010). The SHBG-predicted QRS amplitude component was also associated with lower CV risk (HR = 0.866 per 1-SD increment, p-value=0.004). As higher SHBG corresponds to lower active testosterone, this SHBG-driven attenuation of QRS voltage might suggest a reduction in subclinical myocardial electrical strain or depolarization overload, independent of structural cardiac mass owing to our prior confounder adjustment. Taken together, these findings suggest that androgen-related ECG variation may capture complementary depolarization and repolarization pathways in males.

The menopause-specific analyses highlight another key finding of this study: menopausal status is associated with distinct lead I ECG morphology beyond simple age stratification. Existing literature is highly polarized, focusing either on QTc fluctuations during the menstrual cycle in young females^42^ or on raw ECG changes following hormone therapies in postmenopausal cohorts^43^, rarely bridging both groups under a unified protocol. Our ECG parameter and morphology analysis showed that postmenopausal women exhibit a distinct ECG signature, characterized by higher QRS amplitudes and delayed T-waves, which remained visually apparent across age strata (Figure 2, panel B.2). A plausible physiological explanation is that higher QRS complexes may reflect the higher indexed LVM and LVWT observed in postmenopausal women (Table 1). These structural differences may be related to postmenopausal vascular changes, including reduced oestrogen-mediated vasodilation and increased arterial stiffness^44^. In parallel, delayed T-waves and prolonged QTc intervals are consistent with the postmenopausal hormonal withdrawal, particularly reductions in oestradiol and testosterone, both of which have been linked to QTc prolongation^45,46^. However, these proposed physiological pathways remain speculative based on our observational data, and further mechanistic and longitudinal studies are warranted to elucidate the exact biological drivers behind these menopause-specific ECG shifts.

Given the observational nature of our study, these findings reflect statistical associations rather than definitive causal relationships. Future work leveraging approaches such as Mendelian randomization^47^ could help explore causal pathways and evaluate whether dynamic hormonal changes directly drive electrophysiological remodeling and long-term cardiovascular risk.

### Limitations

A limitation of this work is that the evaluation of CV risk in the female cohort was likely constrained by the lack of sufficient oestradiol data (>80% missingness) and the total absence of progesterone in the UKB database, which prevented a full assessment of the female-specific endocrine-ECG axis and potentially omitted key hormonal drivers of clinical risk that our current models, focused on total testosterone, FAI and SHBG, could not capture. This issue is closely linked to the fact that biological sex and menopausal status were classified based on baseline self-reported data rather than biochemical confirmation via hormonal levels. Furthermore, although a reclassification rule was applied to handle missing baseline menopausal status (13.5%), this strategy could misclassify perimenopausal females or those with atypical hormonal profiles. Although hormone assays represent the clinical gold standard, the automated immunoassay used for oestradiol in the UKB lacks sufficient sensitivity at lower concentrations (minimum reportable concentrations was 175 pmol/L for oestradiol), resulting in undetectable levels for the vast majority of female participants^32^. Additionally, our two-stage regression framework inherently assumes a linear relationship, potentially overlooking non-linear or threshold effects that warrant exploration in future studies. Furthermore, our study was restricted to lead I ECG recordings. While lead I provides robust data for front-plane vector changes, it cannot capture the full spatial and three-dimensional complexity of ventricular depolarization and repolarization, which would be better appreciated through a standard 12-lead ECG.

Finally, while cardiac anatomical confounders, such as indexed LVM and LVWT, were included to account for structural influences on the ECG, these data were exclusively available for the subset of participants who underwent the imaging assessments. Although we implemented interaction terms with the time elapsed between visits to adjust for potential longitudinal structural changes, the inherent variability in cardiac remodelling over time and the reduced sample size in this specific cohort may have introduced residual confounding that could not be fully addressed.

## Conclusion

This study identifies sex- and menopause-dependent associations between hormonal markers and ECG parameters after accounting for clinical, anthropometric, and cardiac anatomical factors that directly translate into long-term CV risk. Androgen-related ECG components, particularly those linked to SHBG and FAI, carry prognostic clinical information with incident CV events primarily in premenopausal females and males. These findings support an endocrine-aware interpretation of ECG parameters and highlight the need for further studies incorporating complete female hormonal profiling and multi-lead ECG morphology.

## Data Availability

Anonymized data and materials generated in this work will be returned to the UK Biobank and can be accessed upon request.

https://www.ukbiobank.ac.uk/use-our-data/apply-for-access/

## Acknowledgements

This research has been conducted using the UK Biobank resource under application number 8256. The authors would like to thank all the participants and professionals contributing to the UK Biobank.

## Funding

This work was supported by project PID2023-148975OB-I00, funded by the Spanish Ministry of Science and Innovation (MCIN/AEI/10.13039/501100011033). PBM and WJY acknowledge the support of the National Institute for Health and Care Research Barts Biomedical Research Centre (NIHR-203330); a delivery partnership of Barts Health NHS Trust, QMUL, St George’s University Hospitals NHS Foundation Trust and St George’s University of London. WJY recognises the National Insitute for Health and Care Research that funded his Academic Clinical Lectureship. This study was conducted using the UK-Biobank [access application-8256], data provided by patients and collected by the NHS as part of their care and support.

## Conflict of interest

All other authors disclose no conflict of interest for this work.

## Notes

### Competing Interest Statement

The authors have declared no competing interest.

