## Supplemental Material for "Sex- and Menopause-Specific Hormone-Related ECG Signatures and Incident Cardiovascular Risk in UK Biobank"

Supplemental Methods

Supplemental Tables

Supplemental Figures

References

**Supplemental Methods**

Definition of cardiovascular events

Diseases were defined by the WHO International Classification of Diseases (ICD) and Related Health Outcomes, Tenth Revision codes, as follows:

| Code | Type | Description |
| --- | --- | --- |
| B332 | ICD10 | Viral carditis |
| G45 | ICD10 | Transient cerebral ischaemic attacks and related syndromes |
| G450 | ICD10 | Vertebro-basilar artery syndrome |
| G453 | ICD10 | Amaurosis fugax |
| G458 | ICD10 | Other transient cerebral ischaemic attacks and related |
| G459 | ICD10 | Transient cerebral ischaemic attack, unspecified |
| I11 | ICD10 | Hypertensive heart disease |
| I110 | ICD10 | Hypertensive heart disease with (congestive) heart failure |
| I119 | ICD10 | Hypertensive heart disease without (congestive) heart failure |
| I130 | ICD10 | Hypertensive heart and renal disease with both (congestive) heart failure |
| I132 | ICD10 | Hypertensive heart and renal disease with both (congestive) heart failure and renal failure |
| I20 | ICD10 | Angina pectoris |
| I200 | ICD10 | Unstable angina |
| I201 | ICD10 | Angina pectoris with documented spasm |
| I208 | ICD10 | Other forms of angina pectoris |
| I209 | ICD10 | Angina pectoris, unspecified |
| I21 | ICD10 | Acute myocardial infarction |
| I210 | ICD10 | Acute transmural myocardial infarction of anterior wall |
| I211 | ICD10 | Acute transmural myocardial infarction of inferior wall |
| I212 | ICD10 | Acute transmural myocardial infarction of other sites |
| I213 | ICD10 | Acute transmural myocardial infarction of unspecified site |
| I214 | ICD10 | Acute subendocardial myocardial infarction |
| I219 | ICD10 | Acute myocardial infarction, unspecified |
| I22 | ICD10 | Subsequent myocardial infarction |
| I220 | ICD10 | Subsequent myocardial infarction of anterior wall |
| I221 | ICD10 | Subsequent myocardial infarction of inferior wall |
| I228 | ICD10 | Subsequent myocardial infarction of other sites |
| I229 | ICD10 | Subsequent myocardial infarction of unspecified site |
| I23 | ICD10 | Certain current complications following acute myocardial infarction |
| I230 | ICD10 | Haemopericardium as current complication following acute myocardial infarction |
| I231 | ICD10 | Atrial septal defect as current complication following acute myocardial infarction |
| I232 | ICD10 | Ventricular septal defect as current complication following acute myocardial infarction |
| I233 | ICD10 | Rupture of cardiac wall without haemopericardium as current complication following acute myocardial infarction |
| I234 | ICD10 | Rupture of chordae tendineae as current complication following acute myocardial infarction |
| I235 | ICD10 | Rupture of papillary muscle as current complication following acute myocardial infarction |
| I236 | ICD10 | Thrombosis of atrium, auricular appendage and ventricle as current complications following acute myocardial infarction |
| I238 | ICD10 | Other current complications following acute myocardial infarction |
| I24 | ICD10 | Other acute ischaemic heart diseases |
| I240 | ICD10 | Coronary thrombosis not resulting in myocardial infarction |
| I248 | ICD10 | Other forms of acute ischaemic heart disease |
| I249 | ICD10 | Acute ischaemic heart disease, unspecified |
| I25 | ICD10 | Chronic ischaemic heart disease |
| I250 | ICD10 | Atherosclerotic cardiovascular disease, so described |
| I251 | ICD10 | Atherosclerotic heart disease |
| I252 | ICD10 | Old myocardial infarction |
| I253 | ICD10 | Aneurysm of heart |
| I254 | ICD10 | Coronary artery aneurysm |
| I255 | ICD10 | Ischaemic cardiomyopathy |
| I256 | ICD10 | Silent myocardial ischaemia |
| I258 | ICD10 | Other forms of chronic ischaemic heart disease |
| I259 | ICD10 | Chronic ischaemic heart disease, unspecified |
| I34 | ICD10 | Nonrheumatic mitral valve disorders |
| I340 | ICD10 | Mitral (valve) insufficiency |
| I341 | ICD10 | Mitral (valve) prolapse |
| I342 | ICD10 | Nonrheumatic mitral (valve) stenosis |
| I348 | ICD10 | Other nonrheumatic mitral valve disorders |
| I349 | ICD10 | Nonrheumatic mitral valve disorder, unspecified |
| I35 | ICD10 | Nonrheumatic aortic valve disorders |
| I350 | ICD10 | Aortic (valve) stenosis |
| I351 | ICD10 | Aortic (valve) insufficiency |
| I352 | ICD10 | Aortic (valve) stenosis with insufficiency |
| I358 | ICD10 | Other aortic valve disorders |
| I359 | ICD10 | Aortic valve disorder, unspecified |
| I36 | ICD10 | Nonrheumatic tricuspid valve disorders |
| I360 | ICD10 | Nonrheumatic tricuspid (valve) stenosis |
| I361 | ICD10 | Nonrheumatic tricuspid (valve) insufficiency |
| I368 | ICD10 | Other nonrheumatic tricuspid valve disorders |
| I369 | ICD10 | Nonrheumatic tricuspid valve disorder, unspecified |
| I37 | ICD10 | Pulmonary valve disorders |
| I370 | ICD10 | Pulmonary valve stenosis |
| I371 | ICD10 | Pulmonary valve insufficiency |
| I372 | ICD10 | Pulmonary valve stenosis with insufficiency |
| I378 | ICD10 | Other pulmonary valve disorders |
| I379 | ICD10 | Pulmonary valve disorder, unspecified |
| I40 | ICD10 | Acute myocarditis |
| I400 | ICD10 | Infective myocarditis |
| I401 | ICD10 | Isolated myocarditis |
| I408 | ICD10 | Other acute myocarditis |
| I409 | ICD10 | Acute myocarditis, unspecified |
| I411 | ICD10 | Myocarditis in viral diseases classified elsewhere |
| I412 | ICD10 | Myocarditis in other infectious and parasitic diseases classified elsewhere |
| I418 | ICD10 | Myocarditis in other diseases classified elsewhere |
| I42 | ICD10 | Cardiomyopathy |
| I420 | ICD10 | Dilated cardiomyopathy |
| I421 | ICD10 | Obstructive hypertrophic cardiomyopathy |
| I422 | ICD10 | Other hypertrophic cardiomyopathy |
| I423 | ICD10 | Endomyocardial (eosinophilic) disease |
| I424 | ICD10 | Endocardial fibroelastosis |
| I425 | ICD10 | Other restrictive cardiomyopathy |
| I426 | ICD10 | Alcoholic cardiomyopathy |
| I427 | ICD10 | Cardiomyopathy due to drugs and other external agents |
| I428 | ICD10 | Other cardiomyopathies |
| I429 | ICD10 | Cardiomyopathy, unspecified |
| I43 | ICD10 | Cardiomyopathy in diseases classified elsewhere |
| I430 | ICD10 | Cardiomyopathy in infectious and parasitic diseases classified elsewhere |
| I431 | ICD10 | Cardiomyopathy in metabolic diseases |
| I432 | ICD10 | Cardiomyopathy in nutritional diseases |
| I438 | ICD10 | Cardiomyopathy in other diseases classified elsewhere |
| I440 | ICD10 | Atrioventricular block, first degree |
| I441 | ICD10 | Atrioventricular block, second degree |
| I442 | ICD10 | Atrioventricular block, complete |
| I447 | ICD10 | Left bundle-branch block, unspecified |
| I460 | ICD10 | Cardiac arrest with successful resuscitation |
| I461 | ICD10 | Sudden cardiac death, so described |
| I469 | ICD10 | Cardiac arrest, unspecified |
| I470 | ICD10 | Reentry ventricular arrhythmia |
| I471 | ICD10 | Supraventricular tachycardia |
| I472 | ICD10 | Ventricular tachycardia |
| I48 | ICD10 | Atrial fibrillation |
| I480 | ICD10 | Paroxysmal atrial fibrillation |
| I481 | ICD10 | Persistent atrial fibrillation |
| I482 | ICD10 | Chronic atrial fibrillation |
| I483 | ICD10 | Typical atrial flutter |
| I484 | ICD10 | Atypical atrial flutter |
| I489 | ICD10 | Atrial fibrillation and atrial flutter, unspecified |
| I490 | ICD10 | Ventricular fibrillation and flutter |
| I499 | ICD10 | Cardiac arrhythmia, unspecified |
| I50 | ICD10 | Heart failure |
| I500 | ICD10 | Congestive heart failure |
| I501 | ICD10 | Left ventricular failure |
| I509 | ICD10 | Heart failure, unspecified |
| I514 | ICD10 | Myocarditis, unspecified |
| I61 | ICD10 | Intracerebral haemorrhage |
| I610 | ICD10 | Intracerebral haemorrhage in hemisphere, subcortical |
| I611 | ICD10 | Intracerebral haemorrhage in hemisphere, cortical |
| I612 | ICD10 | Intracerebral haemorrhage in hemisphere, unspecified |
| I613 | ICD10 | Intracerebral haemorrhage in brain stem |
| I614 | ICD10 | Intracerebral haemorrhage in cerebellum |
| I615 | ICD10 | Intracerebral haemorrhage, intraventricular |
| I616 | ICD10 | Intracerebral haemorrhage, multiple localised |
| I618 | ICD10 | Other intracerebral haemorrhage |
| I619 | ICD10 | Intracerebral haemorrhage, unspecified |
| I63 | ICD10 | Cerebral infarction |
| I630 | ICD10 | Cerebral infarction due to thrombosis of precerebral arteries |
| I631 | ICD10 | Cerebral infarction due to embolism of precerebral arteries |
| I632 | ICD10 | Cerebral infarction due to unspecified occlusion or stenosis of precerebral arteries |
| I633 | ICD10 | Cerebral infarction due to thrombosis of cerebral arteries |
| I634 | ICD10 | Cerebral infarction due to embolism of cerebral arteries |
| I635 | ICD10 | Cerebral infarction due to unspecified occlusion or stenosis of cerebral arteries |
| I638 | ICD10 | Other cerebral infarction |
| I639 | ICD10 | Cerebral infarction, unspecified |
| I64 | ICD10 | Stroke, not specified as haemorrhage or infarction |
| I70 | ICD10 | Atherosclerosis |
| I700 | ICD10 | Atherosclerosis of aorta |
| I7000 | ICD10 | Atherosclerosis of aorta (without gangrene) |
| I7001 | ICD10 | Atherosclerosis of aorta (with gangrene) |
| I702 | ICD10 | Atherosclerosis of arteries of the extremities |
| I7020 | ICD10 | Atherosclerosis of arteries of extremities (without gangrene) |
| I7021 | ICD10 | Atherosclerosis of arteries of extremities (with gangrene) |
| I708 | ICD10 | Atherosclerosis of other arteries |
| I7080 | ICD10 | Atherosclerosis of other arteries (without gangrene) |
| I71 | ICD10 | Aortic aneurysm and dissection |
| I710 | ICD10 | Dissection of aorta [any part] |
| I711 | ICD10 | Thoracic aortic aneurysm, ruptured |
| I712 | ICD10 | Thoracic aortic aneurysm, without mention of rupture |
| I713 | ICD10 | Abdominal aortic aneurysm, ruptured |
| I714 | ICD10 | Abdominal aortic aneurysm, without mention of rupture |
| I715 | ICD10 | Thoracoabdominal aortic aneurysm, ruptured |
| I716 | ICD10 | Thoracoabdominal aortic aneurysm, without mention of rupture |
| I718 | ICD10 | Aortic aneurysm of unspecified site, ruptured |
| I719 | ICD10 | Aortic aneurysm of unspecified site, without mention of rupture |
| I73 | ICD10 | Other peripheral vascular diseases |
| I730 | ICD10 | Raynaud's syndrome |
| I731 | ICD10 | Thromboangiitis obliterans [Buerger] |
| I738 | ICD10 | Other specified peripheral vascular diseases |
| I739 | ICD10 | Peripheral vascular disease, unspecified |
| I74 | ICD10 | Arterial embolism and thrombosis |
| I740 | ICD10 | Embolism and thrombosis of abdominal aorta |
| I741 | ICD10 | Embolism and thrombosis of other and unspecified parts of aorta |
| I742 | ICD10 | Embolism and thrombosis of arteries of the upper extremities |
| I743 | ICD10 | Embolism and thrombosis of arteries of the lower extremities |
| I744 | ICD10 | Embolism and thrombosis of arteries of extremities, unspecified |
| I745 | ICD10 | Embolism and thrombosis of iliac artery |
| I748 | ICD10 | Embolism and thrombosis of other arteries |
| I749 | ICD10 | Embolism and thrombosis of unspecified artery |
| Q20 | ICD10 | Congenital malformations of cardiac chambers and connexions |
| Q200 | ICD10 | Common arterial trunk |
| Q201 | ICD10 | Double outlet right ventricle |
| Q202 | ICD10 | Double outlet left ventricle |
| Q203 | ICD10 | Discordant ventriculoarterial connexion |
| Q204 | ICD10 | Double inlet ventricle |
| Q205 | ICD10 | Discordant atrioventricular connexion |
| Q206 | ICD10 | Isomerism of atrial appendages |
| Q208 | ICD10 | Other congenital malformations of cardiac chambers and connexions |
| Q209 | ICD10 | Congenital malformation of cardiac chambers and connexions, unspecified |
| Q21 | ICD10 | Congenital malformations of cardiac septa |
| Q210 | ICD10 | Ventricular septal defect |
| Q211 | ICD10 | Atrial septal defect |
| Q212 | ICD10 | Atrioventricular septal defect |
| Q213 | ICD10 | Tetralogy of Fallot |
| Q214 | ICD10 | Aortopulmonary septal defect |
| Q218 | ICD10 | Other congenital malformations of cardiac septa |
| Q219 | ICD10 | Congenital malformation of cardiac septum, unspecified |
| Q22 | ICD10 | Congenital malformations of pulmonary and tricuspid valves |
| Q221 | ICD10 | Congenital pulmonary valve stenosis |
| Q222 | ICD10 | Congenital pulmonary valve insufficiency |
| Q224 | ICD10 | Congenital tricuspid stenosis |
| Q225 | ICD10 | Ebstein's anomaly |
| Q228 | ICD10 | Other congenital malformations of tricuspid valve |
| Q229 | ICD10 | Congenital malformation of tricuspid valve, unspecified |
| Q23 | ICD10 | Congenital malformations of aortic and mitral valves |
| Q230 | ICD10 | Congenital stenosis of aortic valve |
| Q231 | ICD10 | Congenital insufficiency of aortic valve |
| Q232 | ICD10 | Congenital mitral stenosis |
| Q233 | ICD10 | Congenital mitral insufficiency |
| Q234 | ICD10 | Hypoplastic left heart syndrome |
| Q238 | ICD10 | Other congenital malformations of aortic and mitral valves |
| Q239 | ICD10 | Congenital malformation of aortic and mitral valves, unspecified |
| Q24 | ICD10 | Other congenital malformations of heart |
| Q240 | ICD10 | Dextrocardia |
| Q241 | ICD10 | Levocardia |
| Q243 | ICD10 | Pulmonary infundibular stenosis |
| Q244 | ICD10 | Congenital subaortic stenosis |
| Q245 | ICD10 | Malformation of coronary vessels |
| Q246 | ICD10 | Congenital heart block |
| Q248 | ICD10 | Other specified congenital malformations of heart |
| Q249 | ICD10 | Congenital malformation of the heart, unspecified |
| 4020 | ICD9 | Hypertensive heart disease, specified as malignant |
| 4021 | ICD9 | Hypertensive heart disease, specified as benign |
| 4029 | ICD9 | Hypertensive heart disease, not specified as malignant or benign |
| 4109 | ICD9 | Acute myocardial infarction |
| 4119 | ICD9 | Other forms of acute ischaemic heart disease |
| 4129 | ICD9 | Old myocardial infarction |
| 4139 | ICD9 | Angina pectoris |
| 4140 | ICD9 | Coronary artherosclerosis |
| 4141 | ICD9 | Aneurysm of heart |
| 4148 | ICD9 | Other specified forms of chronic ischaemic heart disease |
| 4149 | ICD9 | Chronic ischaemic heart disease, unspecified |
| 4229 | ICD9 | Other and unspecified acute myocarditis |
| 4250 | ICD9 | Endomyocardial fibrosis |
| 4251 | ICD9 | Obstructive hypertrophic cardiomyopathy |
| 4252 | ICD9 | Obscure cardiomyopathy of africa |
| 4253 | ICD9 | Endocardial fibroelastosis |
| 4254 | ICD9 | Other primary cardiomyopathies |
| 4255 | ICD9 | Alcoholic cardiomyopathy |
| 4256 | ICD9 | Cardiomyopathy in chagas's diseases |
| 4257 | ICD9 | Nutritional and metabolic cardiomyopathies |
| 4258 | ICD9 | Cardiomyopathy in other diseases classified elsewhere |
| 4259 | ICD9 | Secondary cardiomyopathy, unspecified |
| 4260 | ICD9 | Atrioventricular block, complete |
| 4263 | ICD9 | Other left bundle branch block |
| 4270 | ICD9 | Paroxysmal supraventricular tachycardia |
| 4271 | ICD9 | Paroxysmal ventricular tachycardia |
| 4273 | ICD9 | Atrial fibrillation |
| 4274 | ICD9 | Ventricular fibrillation and flutter |
| 4275 | ICD9 | Cardiac arrest, unspecified |
| 4278 | ICD9 | Other specified cardiac dysrhythmias |
| 4279 | ICD9 | Cardiac dysrhythmias, unspecified |
| 4280 | ICD9 | Congestive heart failure |
| 4281 | ICD9 | Left ventricular failure |
| 4289 | ICD9 | Heart failure, unspecified |
| 4290 | ICD9 | Myocarditis, unspecified |
| 4292 | ICD9 | Cardiovascular disease, unspecified |
| 4319 | ICD9 | Intracerebral haemorrhage |
| 4340 | ICD9 | Cerebral thrombosis |
| 4341 | ICD9 | Cerebral embolism |
| 4349 | ICD9 | Occlusion of cerebral arteries, unspecified |
| 4359 | ICD9 | Transient cerebral ischaemia |
| 4369 | ICD9 | Acute but ill-defined cerebrovascular disease |
| 4400 | ICD9 | Atherosclerosis of aorta |
| 4402 | ICD9 | Atherosclerosis of arteries of the extremities |
| 4408 | ICD9 | Atherosclerosis of other arteries |
| 4410 | ICD9 | Dissection of aorta [any part] |
| 4411 | ICD9 | Thoracic aortic aneurysm, ruptured |
| 4412 | ICD9 | Thoracic aortic aneurysm, without mention of rupture |
| 4413 | ICD9 | Abdominal aortic aneurysm, ruptured |
| 4414 | ICD9 | Abdominal aortic aneurysm, without mention of rupture |
| 4415 | ICD9 | Aortic aneurysm of unspecified site, ruptured |
| 4416 | ICD9 | Aortic aneurysm of unspecified site, without mention of rupture |
| 4430 | ICD9 | Raynaud's syndrome |
| 4431 | ICD9 | Thromboangiitis obliterans [Buerger] |
| 4438 | ICD9 | Other specified peripheral vascular diseases |
| 4439 | ICD9 | Peripheral vascular disease, unspecified |
| 4440 | ICD9 | Embolism and thrombosis of abdominal aorta |
| 4441 | ICD9 | Embolism and thrombosis of other and unspecified parts of aorta |
| 4442 | ICD9 | Embolism and thrombosis of arteries of the extremities |
| 4448 | ICD9 | Embolism and thrombosis of other arteries |
| 4449 | ICD9 | Embolism and thrombosis of unspecified artery |
| 7454 | ICD9 | Ventricular septal defect |
| 7455 | ICD9 | Ostium secundum type atrial septal defect |
| 7461 | ICD9 | Congenital tricuspid stenosis |
| 7462 | ICD9 | Ebstein's anomaly |
| 7463 | ICD9 | Congenital stenosis of aortic valve |
| 7464 | ICD9 | Congenital insufficiency of aortic valve |
| 7465 | ICD9 | Congenital mitral stenosis |
| 7466 | ICD9 | Congenital mitral insufficiency |
| 7467 | ICD9 | Hypoplastic left heart syndrome |
| 74520 | ICD9 | Tetralogy of Fallot |
| 74601 | ICD9 | Congenital pulmonary valve stenosis |
| 74602 | ICD9 | Congenital pulmonary valve insufficiency |
| 74680 | ICD9 | Other specified anomalies of heart (dextrocardia without situs inversus) |
| 74681 | ICD9 | Levocardia |
| 74683 | ICD9 | Pulmonary infundibular stenosis |
| 74687 | ICD9 | Congenital heart block |
| 74688 | ICD9 | Other specified congenital malformations of heart |
| 74699 | ICD9 | Unspecified anomalies of heart |
| K40 | OPCS4 | Saphenous vein graft replacement of coronary artery |
| K401 | OPCS4 | Saphenous vein graft replacement of one coronary artery |
| K402 | OPCS4 | Saphenous vein graft replacement of two coronary arteries |
| K403 | OPCS4 | Saphenous vein graft replacement of three coronary arteries |
| K404 | OPCS4 | Saphenous vein graft replacement of four or more coronary arteries |
| K409 | OPCS4 | Unspecified saphenous vein graft replacement of coronary artery |
| K41 | OPCS4 | Other autograft replacement of coronary artery |
| K411 | OPCS4 | Autograft replacement of one coronary artery NEC |
| K412 | OPCS4 | Autograft replacement of two coronary arteries NEC |
| K413 | OPCS4 | Autograft replacement of three coronary arteries NEC |
| K414 | OPCS4 | Autograft replacement of four or more coronary arteries NEC |
| K42 | OPCS4 | Allograft replacement of coronary artery |
| K424 | OPCS4 | Allograft replacement of four or more coronary arteries |
| K44 | OPCS4 | Other replacement of coronary artery |
| K441 | OPCS4 | Replacement of coronary arteries using multiple methods |
| K442 | OPCS4 | Revision of replacement of coronary artery |
| K449 | OPCS4 | Unspecified other replacement of coronary artery |
| K45 | OPCS4 | Connection of thoracic artery to coronary artery |
| K451 | OPCS4 | Double anastomosis of mammary arteries to coronary arteries |
| K452 | OPCS4 | Double anastomosis of thoracic arteries to coronary arteries NEC |
| K453 | OPCS4 | Anastomosis of mammary artery to left anterior descending coronary artery |
| K454 | OPCS4 | Anastomosis of mammary artery to coronary artery NEC |
| K455 | OPCS4 | Anastomosis of thoracic artery to coronary artery NEC |
| K456 | OPCS4 | Revision of connection of thoracic artery to coronary artery |
| K458 | OPCS4 | Other specified connection of thoracic artery to coronary artery |
| K459 | OPCS4 | Unspecified connection of thoracic artery to coronary artery |
| K49 | OPCS4 | Transluminal balloon angioplasty of coronary artery |
| K491 | OPCS4 | Percutaneous transluminal balloon angioplasty of one coronary artery |
| K492 | OPCS4 | Percutaneous transluminal balloon angioplasty of multiple coronary arteries |
| K493 | OPCS4 | Percutaneous transluminal balloon angioplasty of bypass graft of coronary artery |
| K494 | OPCS4 | Percutaneous transluminal cutting balloon angioplasty of coronary artery |
| K498 | OPCS4 | Other specified transluminal balloon angioplasty of coronary artery |
| K499 | OPCS4 | Unspecified transluminal balloon angioplasty of coronary artery |
| K50 | OPCS4 | Other therapeutic transluminal operations on coronary artery |
| K501 | OPCS4 | Percutaenous transluminal laser coronary angioplasty |
| K502 | OPCS4 | Percutaneous transluminal coronary thrombolysis using streptokinase |
| K503 | OPCS4 | Percutaneous transluminal injection of therapeutic substance into coronary artery NEC |
| K504 | OPCS4 | Percutaenous transluminal atherectomy of coronary artery |
| K508 | OPCS4 | Other specified other therapeutic transluminal operations on coronary artery |
| K509 | OPCS4 | Unspecified other therapeutic transluminal operations on coronary artery |
| K576 | OPCS4 | Percutaneous transluminal ablation of ventricular wall |
| K59 | OPCS4 | Cardioverter defibrillator introduced through vein |
| K591 | OPCS4 | Implantation of cardioverter defibrillator using one electrode lead |
| K592 | OPCS4 | Implantation of cardioverter defibrillator using two electrode leads |
| K593 | OPCS4 | Resiting of lead of cardioverter defibrillator |
| K594 | OPCS4 | Renewal of cardioverter defibrillator |
| K596 | OPCS4 | Implantation of cardioverter defibrillator using three electrode leads |
| K598 | OPCS4 | Other specified cardioverter defibrillator introduced through the vein |
| K599 | OPCS4 | Unspecified cardioverter defibrillator introduced through the vein |
| K607 | OPCS4 | Implantation of intravenous biventricular cardiac pacemaker system |
| K617 | OPCS4 | Implantation of biventricular cardiac pacemaker system |
| K621 | OPCS4 | Percutaneous radiofrequency catheter ablation for atrial fibrillation |
| K622 | OPCS4 | Percutaneous radiofrequency catheter ablation for atrial fibrillation |
| K623 | OPCS4 | Percutaneous radiofrequency catheter ablation for atrial fibrillation |
| K624 | OPCS4 | Percutaneous radiofrequency catheter ablation for atrial fibrillation |
| K641 | OPCS4 | Percutaneous radiofrequency ablation of epicardium |
| K72 | OPCS4 | Other cardioverter defibrillator |
| K721 | OPCS4 | Implantation of subcutaenous cardioverter defibrillator |
| K723 | OPCS4 | Renewal of subcutaneous cardioverter defibrillator |
| K75 | OPCS4 | Percutaneous transluminal balloon angioplasty and insertion of stent into coronary artery |
| K751 | OPCS4 | Percutaneous transluminal balloon angioplasty and insertion of 1-2 drug-eluting stents into coronary artery |
| K752 | OPCS4 | Percutaneous transluminal balloon angioplasty and insertion of 3 or more drug-eluting stents into coronary artery |
| K753 | OPCS4 | Percutaneous transluminal balloon angioplasty and insertion of 1-2 stents into coronary artery |
| K754 | OPCS4 | Percutaneous transluminal balloon angioplasty and insertion of 3 or more stents into coronary artery NEC |
| K758 | OPCS4 | Other specified percutaenous transluminal balloon angioplasty and insertion of stent into coronary artery |
| K759 | OPCS4 | Unspecified percutaenous transluminal balloon angioplasty and insetion of stent into coronary artery |
| X503 | OPCS4 | Advanced cardiac pulmonary resuscitation |
| X504 | OPCS4 | External ventricular defibrillation |
| 1066 | SELFDIAG | Heart/cardiac problem |
| 1067 | SELFDIAG | Peripheral vascular disease |
| 1074 | SELFDIAG | Angina |
| 1075 | SELFDIAG | Heart attack/myocardial infarction |
| 1076 | SELFDIAG | Heart failure/pulmonary odema |
| 1077 | SELFDIAG | Heart arrhythmia |
| 1079 | SELFDIAG | Cardiomyopathy |
| 1081 | SELFDIAG | Stroke |
| 1082 | SELFDIAG | Transient ischaemic attack (tia) |
| 1426 | SELFDIAG | Myocarditis |
| 1471 | SELFDIAG | Atrial fibrillation |
| 1483 | SELFDIAG | Atrial flutter |
| 1485 | SELFDIAG | Irregular heart beat |
| 1487 | SELFDIAG | SVT / supraventricular tachycardia |
| 1488 | SELFDIAG | Mitral valve prolapse |
| 1489 | SELFDIAG | Mitral stenosis |
| 1490 | SELFDIAG | Aortic stenosis |
| 1491 | SELFDIAG | Brain haemorrhage |
| 1492 | SELFDIAG | Aortic aneurysm |
| 1583 | SELFDIAG | Ischaemic stroke |
| 1585 | SELFDIAG | Mitral regurgitation / incompetence |
| 1587 | SELFDIAG | Aortic regurgitation / incompetence |
| 1588 | SELFDIAG | Hypertrophic cardiomyopathy (hcm / hocm) |
| 1589 | SELFDIAG | Pericarditis |
| 1591 | SELFDIAG | Aortic aneurysm rupture |
| 1592 | SELFDIAG | Aortic dissection |
| 1069 | SELFOPER | Heart surgery |
| 1070 | SELFOPER | Coronary angioplasty (ptca) +/- stent |
| 1071 | SELFOPER | Other arterial surgery/revascularisation procedures |
| 1095 | SELFOPER | Coronary artery bypass grafts (cabg) |
| 1097 | SELFOPER | Heart valve surgery |
| 1098 | SELFOPER | Heart transplant |
| 1104 | SELFOPER | Aortic aneurysm/repair or stent |
| 1105 | SELFOPER | Carotid artery surgery/endarterectomy |
| 1107 | SELFOPER | Non-coronary artery angioplasty +/- stent |
| 1108 | SELFOPER | Leg artery angioplasty +/- stent |
| 1109 | SELFOPER | Carotid artery angioplasty +/- stent |
| 1523 | SELFOPER | Triple heart bypass |
| 1548 | SELFOPER | Pacemaker insertion |
| 1549 | SELFOPER | Pacemaker battery change |
| 1550 | SELFOPER | Defibrillator/icd insertion |
| 1551 | SELFOPER | Defibrillator/icd battery change |
| 1553 | SELFOPER | Cardiac ablation |
| 1555 | SELFOPER | Femoral/popliteal/iliac aneurysm repair |

UK Biobank field IDs

For each covariate, we used the most recent measurement obtained prior to or on the date of the ECG acquisition from visit 0 or 1 (prioritizing the earliest available recording).

We considered the following demographic covariates: self-reported sex (UK Biobank field 31), genetic sex (22001), genetic ethnicity, age when attended assessment center (21003), standing height (50), weight (21002), body mass index (BMI, 21001), waist circumference (48) and whole body fat-free mass (23101). We also considered the following functional and life-style covariates: hand grip strength, IPAQ activity group (22032), smoking status (20116) and alcohol drinker status (20117).

Hand grip strength is obtained from the grip strength recorded (46 [left],47 [right]) with the participant's dominant hand (1707). If the participant is ambidextrous, the greater grip strength is considered.

We included the following clinical factors: diastolic blood pressure (DBP, 4079), systolic blood pressure (SBP, 4080), pulse rate (102), diabetes diagnosed by a doctor (2443), total cholesterol (30690), low-density lipoprotein (LDL, 30780) and high-density lipoprotein (HDL, 30760).

Participants from the IMG-UKB cohort (visit 2 and 3) underwent CMR imaging acquisition at rest, allowing us to obtain the following anatomical cardiac measurements: left ventricle (LV) myocardial mass (LVM, 24105), LV myocardial wall thickness (LVWT, 24140) and LV ejection fraction (LVEF, 24103). These parameters were derived from the raw imaging data using automated software, supplemented with custom in-house algorithms previously developed and validated in earlier studies^1^. LVM was indexed to body surface area^1^ using the DuBois formula^2^.

Hormonal assays and derived indices

Participants from the EST-UKB cohort (visit 0 and 1) provided blood samples, allowing us to include in our study the following hormonal measurements: total testosterone (30850) and oestradiol (30800) measured by one step and two step competitive analysis, respectively. We also considered a glycoprotein produced that binds to sex hormones, i.e. sex hormone binding globulin (SHBG) (30830), measured by two step sandwich immunoassay analysis. Serum concentrations of total testosterone, oestradiol and SHBG were measured on the UniCel DXI 800 (Beckman Coulter, Brea, USA).

The oestradiol/testosterone (O/T) ratio was calculated by first converting oestradiol from pmol/L to nmol/L by dividing by 1000, then oestradiol (nmol/L) was divided by testosterone (nmol/L):

$$\frac{O}{T}ratio [a.u.]= \frac{\frac{Oestradiol}{1000} \left( \frac{nmol}{L} \right)}{Total testosterone (\frac{nmol}{L})}$$

The Free Androgen Index (FAI) was calculated by the ratio of total testosterone level to SHBG in percentage:

$$FAI \left( \% \right)=\frac{Total testosterone (\frac{nmol}{L})}{SHBG (\frac{nmol}{L})}*100$$

**Supplemental Table**

| PREMENOPAUSAL FEMALE PARTICIPANTS (N = 980) | | | | |
| --- | --- | --- | --- | --- |
| *ECG Parameter* | **QRS_amplitude_** | | **QT_corrected_** | |
|  | β_norm_ | *p*_i_ | β_norm_ | *p*_i_ |
| Confounding factors | | | | |
| Age | 0.143 | 0.144 | -0.077 | 0.716 |
| BMI | **1** | **<0.001** | **-0.898** | **0.039** |
| WC | **0.725** | **<0.001** | 0.525 | 0.208 |
| Fat-free mass | **-0.277** | **0.040** | **0.597** | **0.043** |
| Hand grip strength | -0.003 | 0.974 | 0.265 | 0.241 |
| Systolic BP | 0.264 | 0.078 | -0.597 | 0.064 |
| Diastolic BP | 0.291 | 0.054 | **1** | **0.002** |
| LDL | 0.117 | 0.222 | **0.570** | **0.006** |
| HDL | -0.167 | 0.086 | 0.177 | 0.400 |
| indexed LVM | -0.068 | 0.850 | -0.387 | 0.619 |
| indexed LVM * ∆t | -0.121 | 0.663 | 0.509 | 0.399 |
| LVEF | -0.096 | 0.354 | -0.139 | 0.534 |
| LVEF * ∆t | 0.022 | 0.911 | 0.317 | 0.458 |
| *ECG Residuals* | **Residual QRS_amplitude_** | | **Residual QT_corrected_** | |
|  | β | *p*_i_ | β | *p*_i_ |
| Hormonal measurements | | | | |
| FAI | **19.170** | **0.038** | 0.377 | 0.620 |

**Supplemental Table 1:** **Association of confounding factors and hormonal measurements with ECG parameters and residuals, respectively, in premenopausal female participants (corresponding to Figure 4, panels A.1 and B.1).** β_norm_ represents the normalized regression coefficient for each ECG model (β/max(|β|)), scaled between -1 and 1 to allow direct comparison, while β is the unnormalized regression coefficient representing the direct effect size on the ECG residuals. p_i_, p-value for each individual predictor; WC, waist circumference; BP, blood pressure; LDL, low-density lipoprotein; HDL, high-density lipoprotein; LVM, left ventricular mass; LVEF, left ventricular ejection fraction; ∆t, time elapsed between hormonal and CMR assessment; FAI, free androgen index. Statistically significant associations are indicated in bold.

| MALE PARTICIPANTS (N = 3,997) | | | | | | | | | | | | | | | | |
| --- | --- | --- | --- | --- | --- | --- | --- | --- | --- | --- | --- | --- | --- | --- | --- | --- |
| *ECG*  *Parameter* | **PR_interval_** | | **QRS_width_** | | **QRS_amplitude_** | | **\|ST_amplitude_\|** | | **QT_c_** | | **T_amplitude_** | | **Tpe_c_** | | **TMV** | |
|  | β_norm_ | *p*_i_ | β_norm_ | *p*_i_ | β_norm_ | *p*_i_ | β_norm_ | *p*_i_ | β_norm_ | *p*_i_ | β_norm_ | *p*_i_ | β_norm_ | *p*_i_ | β_norm_ | *p*_i_ |
| Confounding factors | | | | | | | | | | | | | | | | |
| Age | **0.532** | **<0.001** | 0.097 | 0.542 | **-0.193** | **<0.001** | **-1** | **<0.001** | **0.841** | **<0.001** | **-0.503** | **<0.001** | -0.091 | 0.053 | 0.110 | 0.259 |
| BMI | -0.222 | 0.368 | -0.351 | 0.190 | **1** | **<0.001** | **0.466** | **0.009** | **0.454** | **0.005** | **0.658** | **<0.001** | -0.141 | 0.079 | 0.196 | 0.232 |
| WC | 0.220 | 0.367 | -0.260 | 0.328 | **0.238** | **<0.001** | -0.136 | 0.439 | -0.250 | 0.116 | **-0.195** | **0.019** | 0.099 | 0.211 | 0.206 | 0.206 |
| Fat-free mass | 0.280 | 0.116 | **1** | **<0.001** | **-0.345** | **<0.001** | 0.006 | 0.960 | **0.312** | **0.007** | **-0.172** | **0.005** | -0.080 | 0.167 | -0.057 | 0.629 |
| Hand grip strength | -0.110 | 0.423 | 0.199 | 0.182 | **-0.09** | **0.020** | -0.187 | 0.058 | 0.010 | 0.915 | -0.087 | 0.065 | 0.075 | 0.095 | 0.015 | 0.868 |
| Physical activity | -0.111 | 0.732 | -0.279 | 0.428 | **0.334** | **<0.001** | **0.494** | **0.034** | 0.113 | 0.594 | **0.472** | **<0.001** | -0.207 | 0.050 | -0.381 | 0.078 |
| Current smoker | 0.258 | 0.606 | -0.396 | 0.468 | **-0.316** | **0.025** | 0.235 | 0.514 | -0.300 | 0.359 | **-0.440** | **0.010** | -0.307 | 0.060 | -0.194 | 0.560 |
| Alcohol intake | 0.939 | 0.170 | -0.210 | 0.778 | 0.237 | 0.217 | -0.498 | 0.311 | 0.053 | 0.905 | 0.358 | 0.125 | 0.052 | 0.815 | 0.370 | 0.420 |
| Diabetes | -0.256 | 0.702 | -0.163 | 0.825 | -0.026 | 0.890 | 0.429 | 0.376 | -0.645 | 0.146 | **-1** | **<0.001** | -0.213 | 0.338 | **1** | **0.028** |
| Systolic BP | 0.118 | 0.531 | **0.504** | **0.014** | **0.209** | **<0.001** | **-0.380** | **0.005** | 0.118 | 0.338 | 0.024 | 0.703 | **0.311** | **<0.001** | **0.362** | **0.004** |
| Diastolic BP | -0.105 | 0.581 | **-0.711** | **<0.001** | **0.253** | **<0.001** | 0.073 | 0.596 | **0.264** | **0.033** | **0.132** | **0.042** | **-0.157** | **0.011** | -0.242 | 0.055 |
| Pulse rate | **-1** | **<0.001** | -0.188 | 0.194 | 0.007 | 0.843 | -0.085 | 0.373 | **1** | **<0.001** | **-0.513** | **<0.001** | **1** | **<0.001** | **0.406** | **<0.001** |
| LDL | -0.055 | 0.662 | 0.034 | 0.804 | **0.098** | **0.006** | 0.093 | 0.306 | 0.065 | 0.432 | -0.065 | 0.131 | 0.029 | 0.485 | 0.090 | 0.285 |
| HDL | 0.114 | 0.380 | 0.034 | 0.808 | **-0.065** | **0.075** | 0.042 | 0.649 | 0.107 | 0.209 | 0.079 | 0.074 | **-0.089** | **0.035** | 0.033 | 0.702 |
| indexed LVM | **0.906** | **0.017** | 0.652 | 0.112 | **0.297** | **0.005** | **0.841** | **0.002** | 0.134 | 0.586 | 0.049 | 0.700 | 0.103 | 0.401 | 0.161 | 0.524 |
| indexed LVM * ∆t | -0.603 | 0.060 | -0.673 | 0.052 | **-0.192** | **0.032** | **-0.593** | **0.010** | -0.031 | 0.881 | -0.065 | 0.549 | -0.008 | 0.941 | -0.078 | 0.717 |
| LVEF | **-0.635** | **<0.001** | **-0.429** | **0.003** | **-0.093** | **0.012** | **-0.266** | **0.005** | -0.040 | 0.645 | -0.033 | 0.461 | 0.013 | 0.767 | 0.141 | 0.112 |
| LVEF * ∆t | **0.632** | **0.004** | 0.358 | 0.135 | **0.123** | **0.046** | **0.491** | **0.002** | 0.041 | 0.778 | 0.010 | 0.891 | -0.022 | 0.757 | -0.055 | 0.710 |
| *ECG*  *Residuals* | **Residual PR_interval_** | | **Residual QRS_width_** | | **Residual**  **QRS_amplitude_** | | **Residual**  **\|ST_amplitude_\|** | | **Residual**  **QT_c_** | | **Residual**  **T_amplitude_** | | **Residual Tpe_c_** | | **Residual TMV** | |
| Hormonal measurements | | | | | | | | | | | | | | | | |
|  | β | *p*_i_ | β | *p*_i_ | β | *p*_i_ | β | *p*_i_ | β | *p*_i_ | β | *p*_i_ | β | *p*_i_ | β | *p*_i_ |
| SHBG | 0.026 | 0.954 | 0.174 | 0.520 | **-13.19** | **0.010** | -0.595 | 0.073 | **0.833** | **0.014** | -1.847 | 0.223 | 0.159 | 0.303 | 0.021 | 0.179 |
| FAI | -0.002 | 0.995 | -0.125 | 0.644 | 0.936 | 0.856 | **1.126** | **<0.001** | **-0.951** | **0.005** | 2.841 | 0.061 | -0.127 | 0.410 | -0.030 | 0.056 |

**Supplemental Table 2:** **Association of confounding factors and hormonal measurements with ECG parameters and residuals, respectively, in male participants (corresponding to Figure 4, panels A.2 and B.2).** β_norm_ represents the normalized regression coefficient for each ECG model (β/max(|β|)), scaled between -1 and 1 to allow direct comparison, while β is the unnormalized regression coefficient representing the direct effect size on the ECG residuals. *p*_i_, p-value for each individual predictor; WC, waist circumference; BP, blood pressure; LDL, low-density lipoprotein; HDL, high-density lipoprotein; LVM, left ventricular mass; LVEF, left ventricular ejection fraction; ∆t, time elapsed between hormonal and CMR assessment; SHBG, sex hormone-binding globulin; FAI, free androgen index. Statistically significant associations are indicated in bold.

|  | **Premenopausal females (N = 980)** | |
| --- | --- | --- |
| CV event (count, %)  Control (count, %)  Median follow-up (years) | 20 (2.04%)  960 (97.96%)  12.9 | |
|  | HR (95% CI) | P-value |
| QRS_amp_ residual explained  by FAI (per 1-SD increment) | 1.417  (1.055 – 1.905) | **0.020** |
|  | **Males (N = 3,997)** | |
| CV event (count, %)  Control (count, %)  Median follow-up (years) | 335 (8.4%)  3,662 (91.6%)  12.8 | |
|  | HR (95% CI) | P-value |
| QRS_amp_ residual explained  by SHBG (per 1-SD increment) | 0.866  (0.784 – 0.956) | **0.004** |
| QT_c_ residual explained  by SHBG (per 1-SD increment) | 1.158  (1.048 – 1.279) | **0.004** |
| \|ST_amp_\| residual explained  by FAI (per 1-SD increment) | 0.760  (0.665 – 0.868) | **<.0001** |
| QT_c_ residual explained  by FAI (per 1-SD increment) | 1.317  (1.152 – 1.506) | **<.0001** |

**Supplemental Table 3: Cox proportional hazards models for cardiovascular risk associated with hormone-related ECG components, graphically represented in Figure 4 (panels C).** Premenopausal females- and males-specific analyses are shown in panels C.1 and C.2, respectively. HR: Hazard Ratio; CI: Confidence Interval; SHBG: Sex Hormone-Binding Globulin; FAI: Free Androgen Index; QRS_amp_: QRS complex amplitude; |ST_amp_|: Absolute ST-segment amplitude; QT_c_: Corrected QT interval. Bold p-values indicate statistical significance at the p-value < 0.05.

**Supplementary Figures**

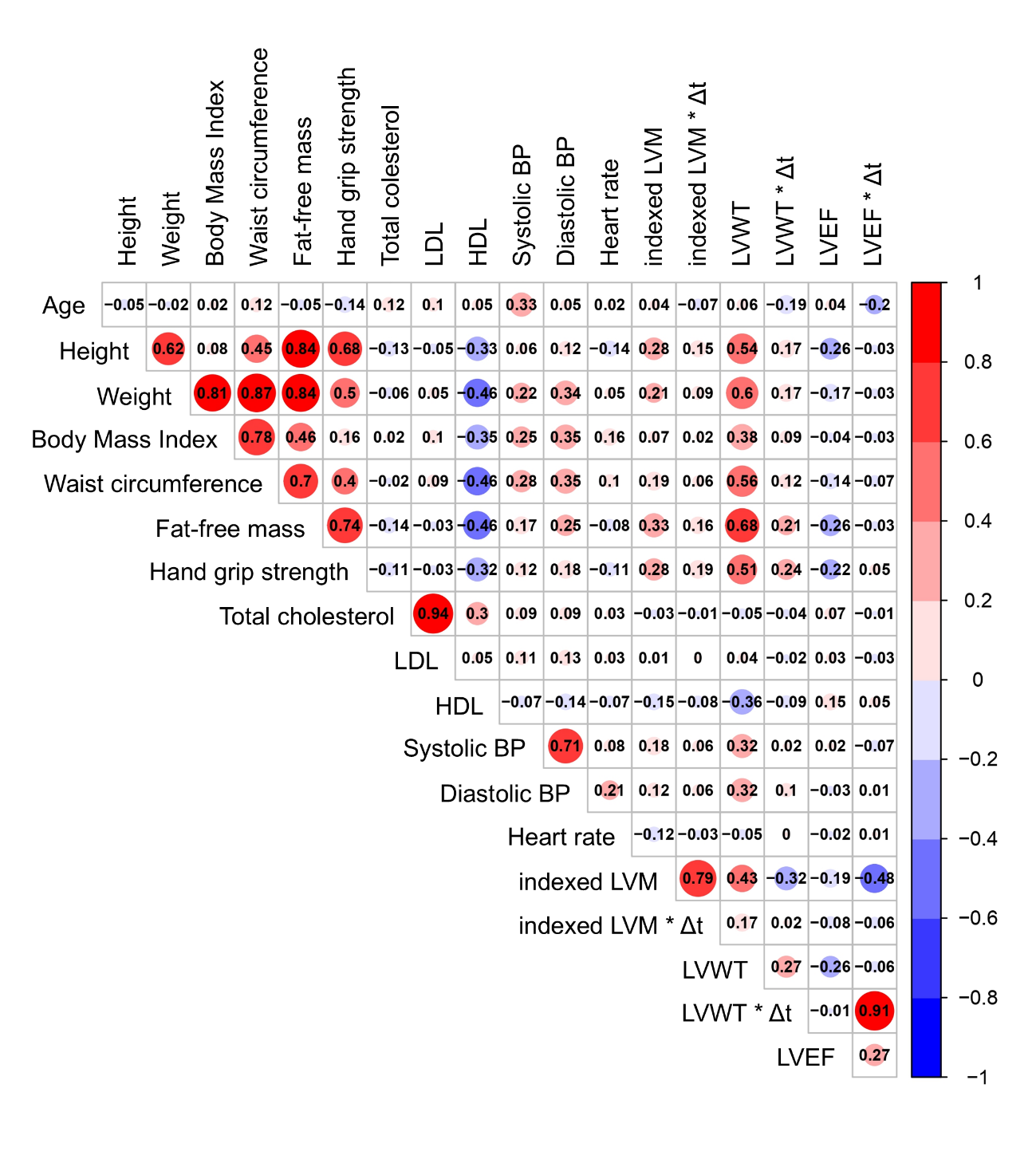

**Supplemental Figure 1: Multicollinearity assessment of continuous covariates.** Spearman rank correlation matrix for continuous covariates considered in the adjustment models. Circle size and color intensity indicate the magnitude of the correlation coefficient. Variable pairs with |r| > 0.80 were considered highly collinear, and one variable was excluded based on physiological relevance and model interpretability. LDL, Low-density Lipoprotein; HDL, High-density Lipoprotein; BP, Blood Pressure; LVM, Left Ventricle Mass; LVWT, Left Ventricle Wall Thickness; LVEF, Left Ventricle Ejection Fraction; ∆t, time elapsed between visits.

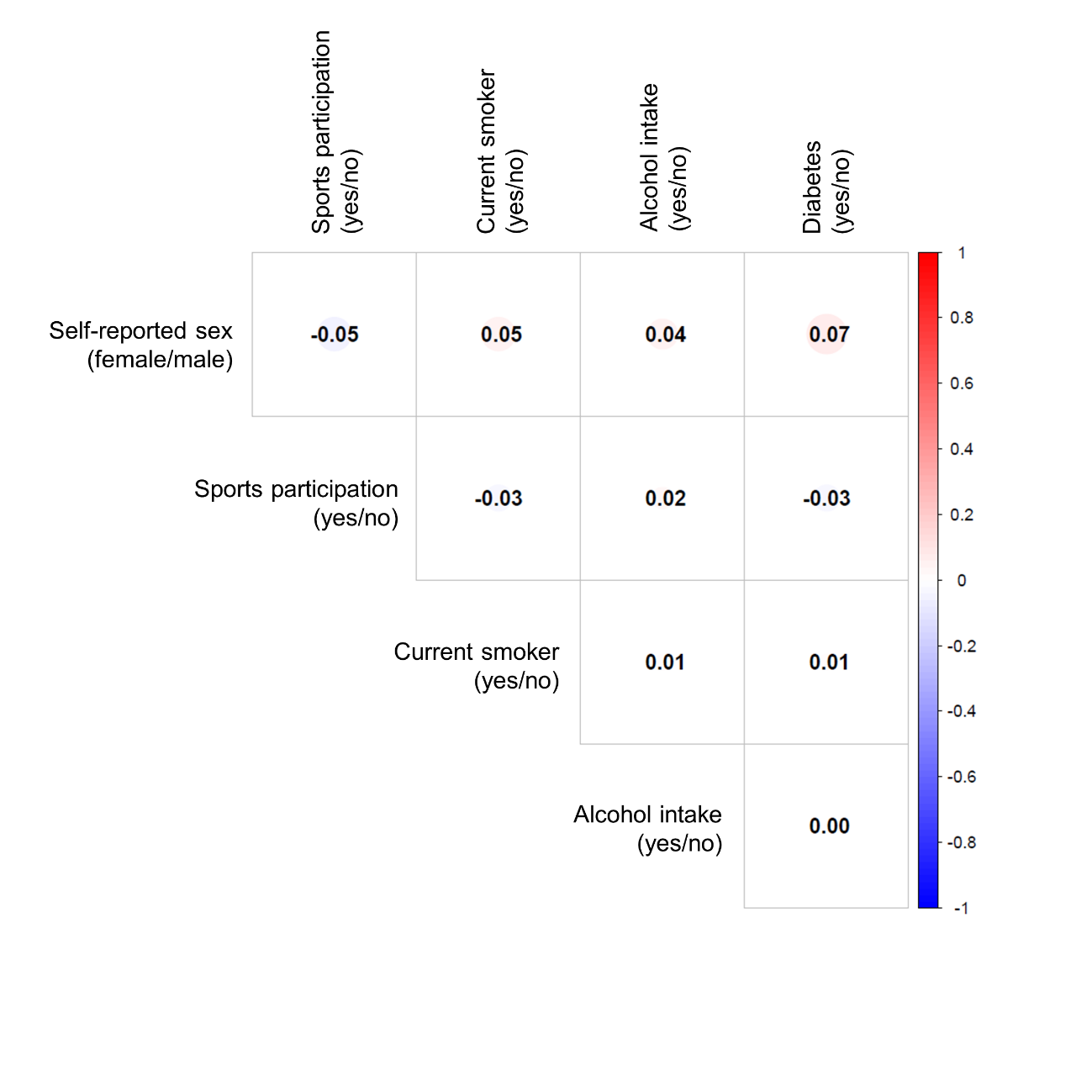

**Supplemental Figure 2: Multicollinearity assessment of categorical covariates.** Phi correlation matrix for categorical covariates considered in the adjustment models. Circle size and color intensity indicate the magnitude of the correlation coefficient. Variables pairs with |r| > 0.80 were considered highly collinear.

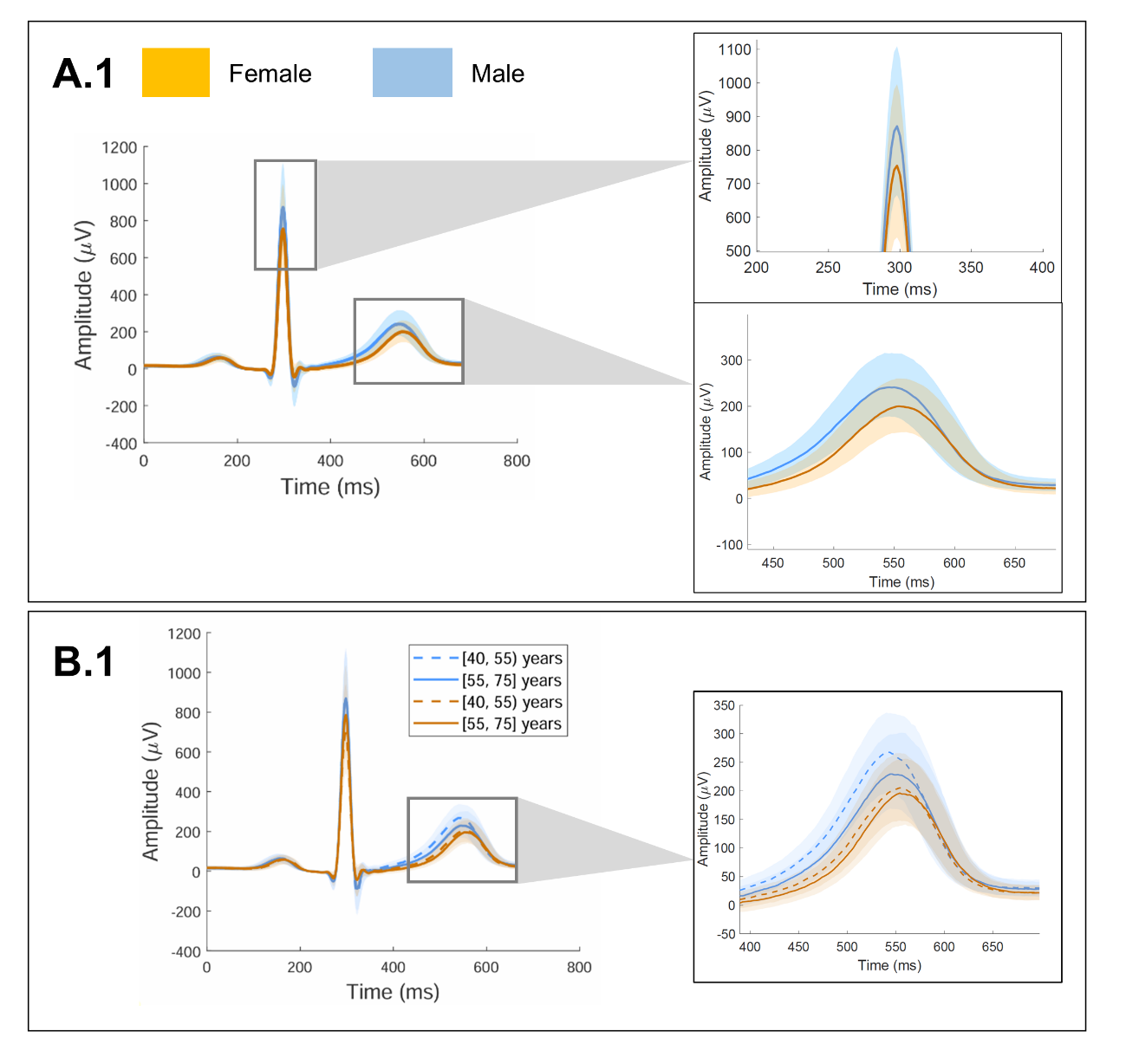

**Supplemental Figure 3: High-magnification views of sex-specific lead I ECG morphology.** High-magnification views of the QRS complex and T-wave from the median lead I heartbeat morphologies shown in Figure 2B. Shaded bands represent the 25^th^-75^th^ percentile range of aligned individual median beats. Panels show the total cohort (A.1) and age-stratified comparisons (B.1).

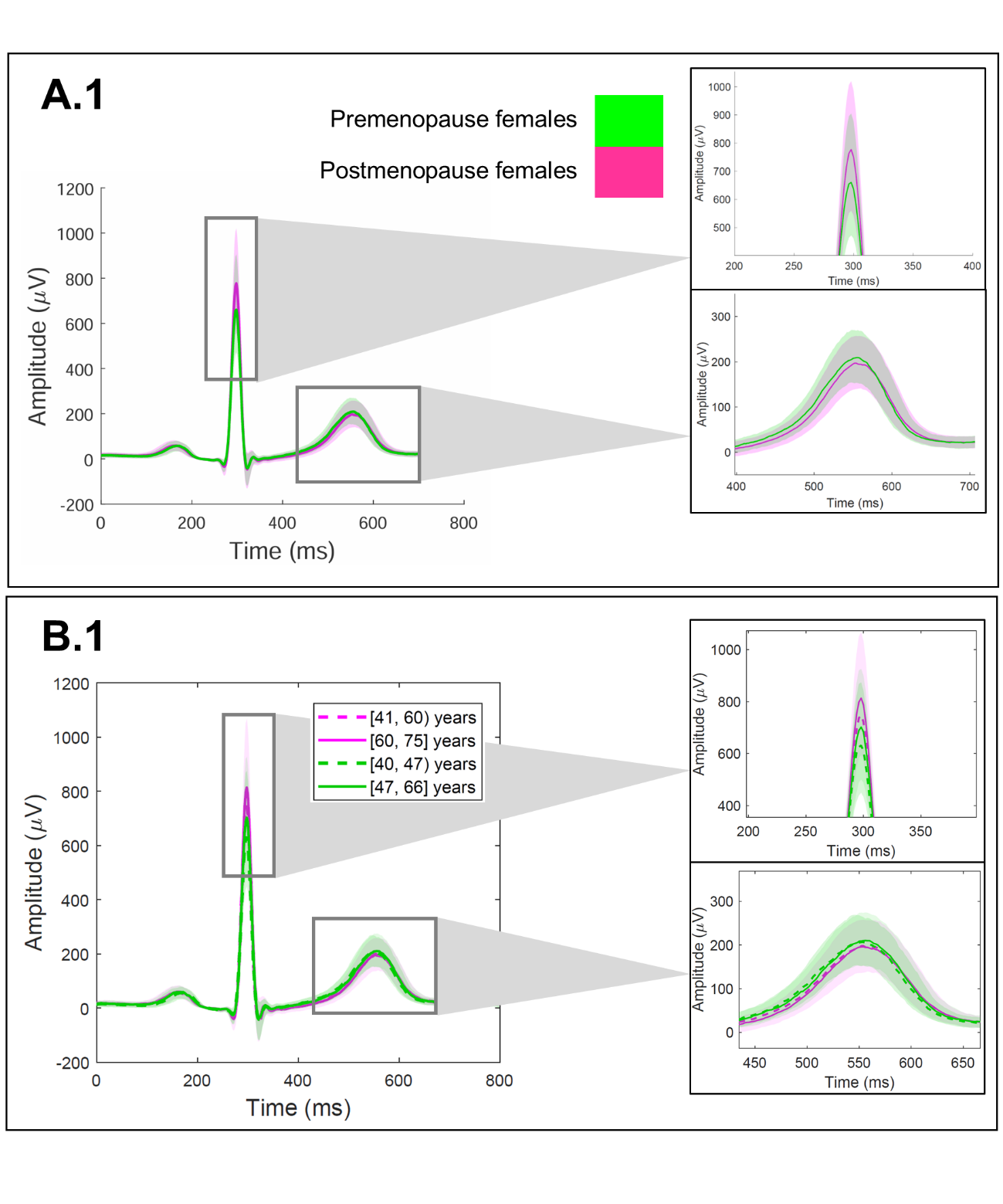
**Supplemental Figure 4: High-magnification views of menopause-specific lead I ECG morphology.** High-magnification views of the QRS complex and T-wave from the median lead I heartbeat morphologies shown in Figure 2 (B.2), stratified by menopausal status. Shaded bands represent the 25^th^-75^th^ percentile range of aligned individual median beats. Panels show the total female cohort (A.1) and age-stratified comparisons (B.1).
